# Pangenome discovery of missing autism variants

**DOI:** 10.1101/2025.07.21.25331932

**Authors:** Yang Sui, Jiadong Lin, Michelle D. Noyes, Youngjun Kwon, Isaac Wong, Nidhi Koundinya, William T. Harvey, Mei Wu, Kendra Hoekzema, Katherine M. Munson, Gage H. Garcia, Jordan Knuth, Julie Wertz, Tianyun Wang, Kelsey Hennick, Druha Karunakaran, Rafael A. Polo Prieto, Rebecca Meyer-Schuman, Fisher Cherry, Davut Pehlivan, Bernhard Suter, Jonas A. Gustafson, Danny E. Miller, Human Pangenome Reference Consortium (HPRC), Hanna Berk-Rauch, Tomasz J. Nowakowski, Aravinda Chakravarti, Huda Y. Zoghbi, Evan E. Eichler

**Author notes:** **Corresponding author:** Evan E. Eichler, Ph.D. Department of Genome Sciences, University of Washington School of Medicine 3720 15th Ave NE, S413C, Box 355065, Seattle, WA 98195-5065.

## Abstract

Autism spectrum disorders (ASDs) are genetically and phenotypically heterogeneous and the majority of cases still remain genetically unresolved. To better understand large-effect pathogenic variation, we generated long-read sequencing data to construct phased and near-complete genome assemblies (average contig N50=43 Mbp, QV=56) for 189 individuals from 51 families with unsolved cases of autism. We applied read- and assembly-based strategies to facilitate comprehensive characterization of *de novo* mutations (DNMs), structural variants (SVs), and DNA methylation profiles. Merging common SVs obtained from long-read pangenome controls, we efficiently filtered >97% of common SVs exclusive to 87 offspring. We find no evidence of increased autosomal SV burden for probands when compared to unaffected siblings yet note a trend for an increase of SV burden on the X chromosome among affected females. We establish a workflow to prioritize potential pathogenic variants by integrating autism risk genes and putative noncoding regulatory elements defined from ATAC-seq and CUT&Tag data from the developing cortex. In total, we identified three pathogenic variants in *TBL1XR1*, *MECP2*, and *SYNGAP1*, as well as nine candidate *de novo* and biparental homozygous SVs, most of which were missed by short-read sequencing. Our work highlights the potential of phased genomes to discover complex more pathogenic mutations and the power of the pangenome to restrict the focus on an increasingly smaller number of SVs for clinical evaluation.

## INTRODUCTION

Autism spectrum disorder (ASD) is a class of neurodevelopmental disorders (NDDs) characterized by challenges in social interaction, communication, and repetitive behaviors, with symptoms and severity varying widely among individuals. Currently, the median prevalence of ASD globally is approximately 1%, with a male-to-female ratio of about 4 to 1 (Zeidan et al. 2022). Genetic investigations into rare variants have typically focused on gene-disruptive *de novo* mutations (DNMs), rare inherited variants, and copy number variants (CNVs) discovered through meta-analyses of short-read sequencing (SRS) and earlier microarray data. These variants account for an estimated 20% of ASD cases and have led to the discovery of ∼1000 risk genes and numerous CNVs associated with NDDs (Zhou et al. 2022; Wilfert et al. 2021; Coe et al. 2019; Wang et al. 2022; Fu et al. 2022; Satterstrom et al. 2020; Yoon et al. 2021; Sebat et al. 2007). The underlying mutations for the remaining cases, including structural variants (SVs) mapping to repetitive regions, remain poorly understood.

SVs, including deletions (DEL), insertions (INS), inversions, translocations, large-scale CNVsand other complex rearrangements, are defined as affecting ≥50 base pairs of DNA. They have been shown to have larger effects (Sudmant, Rausch, et al. 2015; Scott et al. 2021) because they can disrupt coding or noncoding regulatory regions, alongside protein-coding genes, thereby playing a critical role in gene regulation and human disease (Olivucci et al. 2024; Carvalho and Lupski 2016; Collins and Talkowski 2025). However, many SVs occur in technically and methodologically challenging regions, particularly repetitive sequences, making them difficult to detect and completely characterize using conventional SRS approaches such as Illumina whole-genome sequencing (WGS) or whole-exome sequencing (WES). Similarly, a subset of smaller single-nucleotide variants (SNVs) and indels can be missed by SRS because of their association with low-complexity and repetitive DNA (Fu et al. 2022; Wilfert et al. 2021; Wang et al. 2022; Satterstrom et al. 2020).

Long-read sequencing (LRS) data (15-30 kbp) significantly enhances the sensitivity of variant detection, especially in repetitive DNA (Chaisson et al. 2019). Recent studies have revealed that LRS data provide access to ∼91% of the human genome, substantially increasing DNM discovery by ∼30% and SV discovery by over 47% when compared to SRS datasets (Logsdon et al. 2020; Noyes et al. 2022; Ebert et al. 2021; Zhao et al. 2021; Collins and Talkowski 2025; Wojcik et al. 2023; Mastrorosa et al. 2023). Consequently, LRS has been increasingly applied to a variety of unsolved patients and disorders to enhance pathogenic variant discovery, although most studies to date have involved relatively modest cohorts focused almost entirely on read-based discovery. For example, Hiatt et al. reanalyzed 10 NDD families and 86 probands using Pacific Biosciences (PacBio) high-fidelity (HiFi) LRS and found an additional yield of 7.3% beyond SRS mainly in the coding regions. Sanchis-Juan et al. applied Oxford Nanopore Technologies (ONT) LRS to complement SRS in four probands (Hiatt et al. 2024; 2021; Sanchis-Juan et al. 2023; Pauper et al. 2021). Moreover, the ability to accurately call methylated CpGs, especially from ONT LRS, has the added benefit of simultaneously discovering potential imprinting defects (Paschal et al. 2025).

Beyond read-based variant discovery, the combination of LRS technologies (ONT and PacBio) has facilitated the construction of near-complete telomere-to-telomere (T2T) genome assemblies as part of the Human Genome Structural Variation Consortium (HGSVC) and the Human Pangenome Reference Consortium (HPRC) (Ebert et al. 2021; Logsdon, Rozanski, et al. 2024; Liao et al. 2023). These consortia recently made hundreds of diverse human genomes publicly available. This resource is potentially valuable to the clinical genetics community because variant discovery is essentially complete providing a control to assess the frequency of variants in regions typically unassayable by SRS and therefore absent or unreliable in associated databases such as gnomAD (Collins et al. 2020; S. Chen et al. 2024). Moreover, assembly-based comparisons between offspring and parental genomes have been shown to further increase the power to discover DNM by essentially eliminating reference biases (Porubsky et al, 2025).

Using LRS assembly approaches, we sought to construct reference-free genome assemblies comparable to the HGSVC and HPRC controls for all members of autism families. In this study, we present our initial LRS and assembly resource of 189 individuals from 51 unsolved ASD families where no pathogenic variant was previously identified in the proband via conventional Illumina SRS. To build reference resources, we constructed near-complete genomes for each individual and assigned a workflow to systematically identify variants from high-quality assemblies; we then compared with HPRC and HGSVC population controls to discover and validate *de novo* and ultra-rare variants as new candidates for ASD. Importantly, constructing genomes comparable to pangenome references, allowed us to dramatically reduce the variant search space for SVs highlighting the increasing utility of the pangenome for disease variant discovery.

## RESULTS

### Sequence and assembly of genomes from unsolved autism families

We focused on the sequence and assembly of genomes from 51 unsolved simplex ASD families. The set included 46 families (174 individuals) with idiopathic autism from the Simons Simplex Collection (SSC) and Study of Autism Genetics Exploration (SAGE) and five families with a diagnosis of Rett syndrome (15 individuals) (**Fig. 1a, Dataset S1**). The Rett families had been previously screened using either gene panels or WES with no MECP2 pathogenic mutation reported by clinical testing labs after multiple attempts (**Dataset S1, Methods**). Similarly, the 46 families were part of large-scale CNV and WGS initiatives over the last decade where no pathogenic variant had been reported by multiple groups, including our own (Coe et al. 2019; Sanders et al. 2011; Wilfert et al. 2021; Turner et al. 2017). For 36 families, there was also an unaffected sibling serving as a genetic control (17 of these quads were sex-matched).

**Figure 1.**
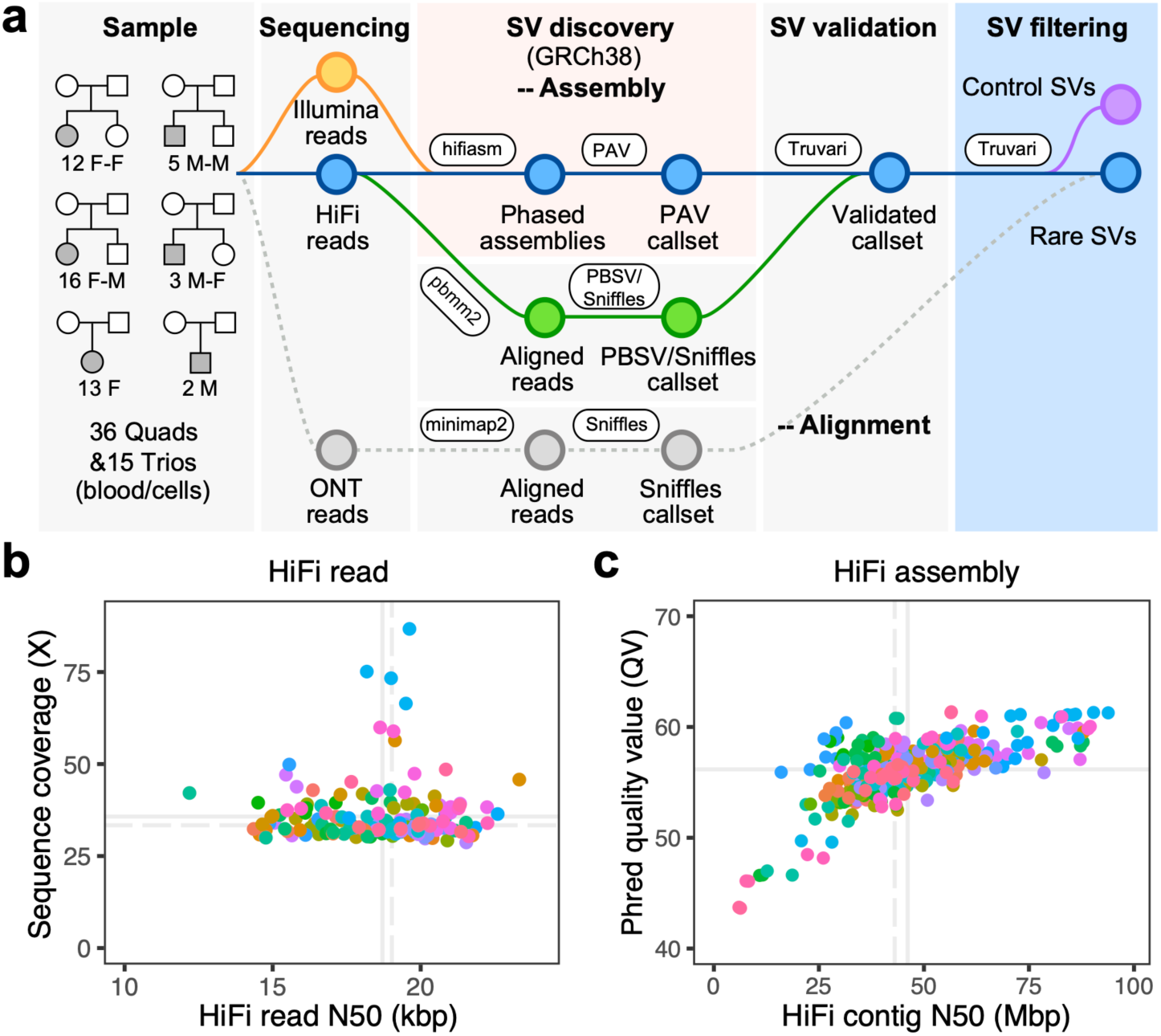
Long-read analysis sequencing and assembly. **a)** Schematic workflow of LRS data generation and SV discovery with pedigree structures of the 51 unsolved ASD families (F=female; M=male). LRS data (PacBio HiFi and ONT) and phased genomes were constructed using hifiasm; SVs were discovered by PAV and validated via Truvari with read-based callers, PBSV and Sniffles (analyses tools indicated in oval boxes). Validated SVs were filtered using a pangenome of 108 control genomes from the HPRC and HGSVC to define a rare SV callset private to the autism families (**Dataset S1**). **b)** HiFi reads N50 and genomic coverage per sample (members of the same family are color coded). **c)** Sequence accuracy (QV) and contig N50 length for each HiFi-phased genome assembly. Solid lines represent mean values, while dashed lines indicate median values.

We developed an LRS workflow to enhance variant discovery using reference-level quality genomes with a particular emphasis on characterizing previously undetected SVs and DNMs. First, we sequenced all 189 genomes using PacBio HiFi sequencing technology from peripheral blood (n=139), cell lines (n=16), or a mixture of both (n=34) when DNA from peripheral lymphocytes was limited. Per sample, we generated an average of 36-fold sequence coverage with an overall N50 length of 19 kbp after extensive quality control (**Fig. 1b, Dataset S1, Methods, Fig. S1**). Using parental Illumina reads and hifiasm (Cheng et al. 2021), we generated haplotype-resolved genome assemblies for each of the 87 offspring. The resulting assemblies are highly contiguous (average contig N50 of 43 Mbp (**Fig. 1c**)) and highly accurate (QV=56). The assembly quality is comparable to that of control pangenomes (with mean QV of 56) from the HPRC (Liao et al. 2023) and HGSVC (Logsdon, Rozanski, et al. 2024; Ebert et al. 2021).

### Variant discovery

For each family, SNVs and small indel callsets were generated by the GATK (Poplin, Ruano-Rubio, et al. 2018) and DeepVariant (Poplin, Chang, et al. 2018) callers using T2T-CHM13v2.0-aligned HiFi reads mainly from blood samples (n=172). Putative DNMs were further validated by confirming their presence in ONT (**Fig. S2**) and Illumina data, as previously described (Porubsky, Dashnow, et al. 2025; Noyes et al. 2025). We discovered on average 96 DNMs per child (n=78), with approximately 83% predicted to be germline and the remainder arising postzygotically. SVs in each individual were identified from the phased assemblies using PAV (Ebert et al. 2021) with GRCh38 as the reference genome. GRCh38 was selected due to the broad availability of annotation resources mapped to this assembly, many of which were integrated into our analyses to facilitate comprehensive functional characterization of putative disruptive variants. Each SV was considered validated if supported by at least one of the alignment-based SV callers, either PacBio structural variant caller (PBSV), Sniffles (Smolka et al. 2024), or both (**Methods**). We aggregated the validated SVs from all 189 study samples and compared them with SVs observed in the 108 population controls from the HPRC and HGSVC via Truvari (English et al. 2022), focusing on rare SVs exclusive to ASD families.

Based on PAV analysis of each child’s assembly, we identified an average of 27,576 SVs per diploid genome of which 20,716 ± 9 (95% confidence limits) SVs were also supported by alignment-based methods (**Fig. 2a**). We note that two Rett-like samples where DNA was limited (HYZ204_p1 and HYZ207_p1) have lower contig N50 and lower assembly QV, resulting in fewer validated SVs (**Fig. 2a, Fig. S3**). The average Mendelian concordance rate for SVs across 87 trios is 90.4%. This high-confidence SV callset, thus, affects 11,074,300 ± 4,273 bp (95% confidence limits) per sample (0.4% of the genome). We applied the same SV discovery approach to 108 pangenome controls resulting in on average 24,341 ± 36 validated SVs per diploid genome (**Fig. S3**). This approach improved the sensitivity of control SV detection and facilitated more effective filtering of SVs in the sample set. Both pangenome controls and study samples are from diverse superpopulations. And, as expected, genomically diverse African samples had higher numbers of SVs than non-Africans (**Fig. S3**). We integrated the two callsets for a total of 271,375 nonredundant SVs and observed an expected SV size distribution with modes at 300 bp and 6 kbp, corresponding to Alu and LINE retrotransposition events, respectively (**Fig. S4**). After filtering with the pangenome we identified a total of 33,548 nonredundant SVs (57,716 genotyped SVs, **Dataset S2**) that were exclusively observed in the 87 children (51 probands and 36 unaffected siblings). At the individual sample level, we effectively filtered ∼97% of SVs per child, resulting in approximately 663 rare SVs per child for further consideration (affected vs. unaffected, Z = -0.23, p=0.82, Mann-Whitney U Test, **Fig. 2a**). After testing for Mendelian inheritance, the autism set was further reduced to 25,272 nonredundant SVs. We then classified rare SVs into six categories (**Fig. 2b**) including autosomal heterozygous (n=33,701), autosomal homozygous (n=6,639), X chromosome heterozygous (n=1,129), X chromosome homozygous (n=277), and hemizygous events from males on the X (n=565) and Y (n=141) chromosomes. We evaluated the remaining low-confidence SV calls through a suite of tools that test for transmission and *de novo* variants (**Methods**). We validated a total of 46 *de novo* SVs in 51 probands (n=26) and 36 siblings (n=20) that were absent from 108 controls (affected vs. unaffected, p=0.84, OR=0.92, χ² test).

**Figure 2.**
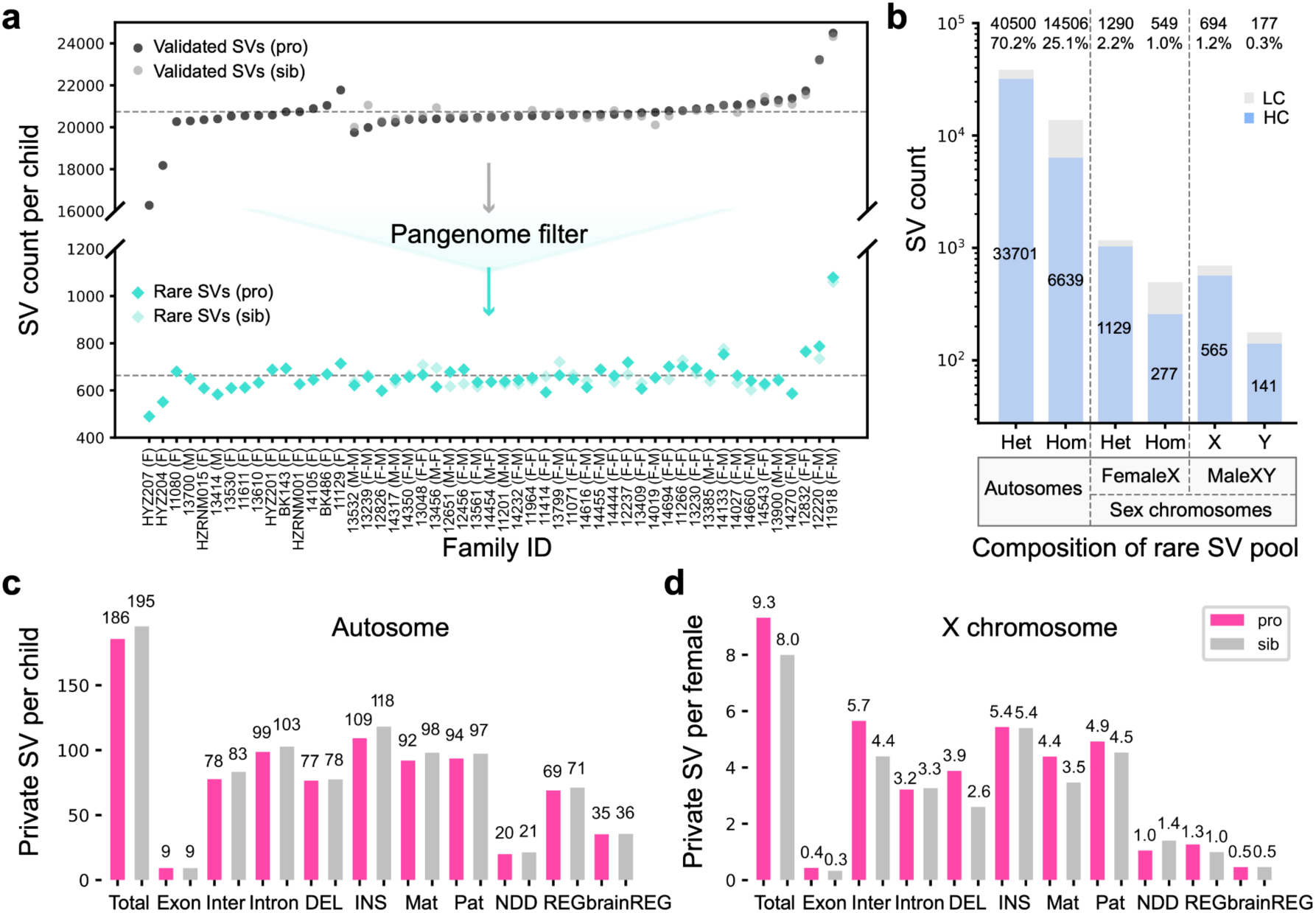
SV discovery, filtering and burden in ASD families. **a)** SV discovery in probands (dark) and unaffected sibling (light) before (top) and after (bottom) pangenome filtering for 51 families with idiopathic autism. Proband sex versus unaffected sibling shown in parenthesis after family IDs. **b)** High-(HC) and low-confidence (LC) SVs by genotype class for autosomes and sex chromosomes. Het: heterozygous SVs. Hom: homozygous SVs. HC: high-confidence SVs confirmed by Mendelian inheritance of parental SV calls. LC: low-confidence SVs that initially deviated from Mendelian inheritance patterns in the collapsed table but were subsequently curated through further evaluation. The histogram compares the **c)** autosomal private SV burden and **d)** X chromosome burden (females only) between probands (pink) and unaffected siblings (gray) for 51 probands (41 females and 10 males) and 36 unaffected siblings (15 females and 21 males). Different functional categories of SV classes are considered: protein-coding and UTR (Exon), intergenic (Inter), intronic (Intron), deletions (DEL), insertions (INS), paternally inherited (Pat), maternally inherited (Mat), those overlapping neurodevelopmental genes (NDD), brain-derived regulatory regions (brainREG) or regulatory regions more generally (REG). No significant differences (χ² test p-values exceeding 0.05) in the number of SVs between probands and siblings were observed across these categories, with a trend observed on the X chromosome for enrichment of SVs on affected females compared to unaffected sisters.

### SV burden analyses

Because the majority of rare SVs (96%) map to noncoding DNA, we annotated all SVs for regulatory potential as well as association with known NDD genes. To define putative regulatory elements (REG), we integrated published datasets from ENCODE with regulatory sequences predicted from single-cell and bulk ATAC-seq and CUT&Tag experiments performed on developmentally staged material (16-24 weeks) from the cerebral cortex (**Methods**). The brain-derived regulatory regions (brainREG) annotation resulted in a 45% increase in the total number of regulatory SVs (**Fig. 2c-d**). For NDD candidate genes, we primarily focused on those previously reported by SFARI, Fu et al., Zhou et al., Satterstrom et al. and Wang et al. (Zhou et al. 2022; Fu et al. 2022; Wang et al. 2022; Satterstrom et al. 2020). We considered three classes of SVs, namely: *de novo*, homozygous, and private inherited SVs defined as those observed only once in the parental population (Wilfert et al. 2021). On autosomes, we identified 9,475 private SVs in 51 probands and 7,035 private SVs in 36 unaffected siblings. The rates of autosomal SVs (**Fig. 2c, Fig. S5**) do not differ significantly between probands and unaffected siblings across various categories (nominal p > 0.05, OR < 1, χ² test). For X chromosome SVs, we analyzed males and females separately. Female probands exhibited a slight excess of X chromosome private SVs compared to the unaffected female siblings (nominal p = 0.29, OR = 1.16, adjusted p = 1, χ² test), particularly for deletions (**Fig. 2d**, nominal p = 0.09, OR = 1.49, adjusted p = 1, χ² test). We also considered 1,417 private homozygous SVs that were inherited from two heterozygous parents on autosomes in only one family but were never observed as homozygous in any controls. Once again, we observed a slight enrichment in the number of these private biparental homozygous SVs in probands relative to unaffected siblings (**Fig. S6**, p > 0.05, χ² test).

### Sex chromosome assembly analyses

In addition to SVs, we constructed nearly complete X and Y chromosomes by leveraging LRS and parental sequence data. Excluding the pseudoautosomal region (PAR), centromere and extremely repetitive Yq12 heterochromatic regions, we estimate that on average 96% of the X chromosome and 75% of the Y chromosome (**Figs. 3a-b, Fig. S7**) can be aligned (**Methods**). The assemblies served two purposes: they validated DNMs on the sex chromosomes and allowed parental transmission to be fully assessed (i.e., paternal Y and maternal vs. paternal X chromosomes) without use of a reference genome (**Fig. 3c-d**).

**Figure 3.**
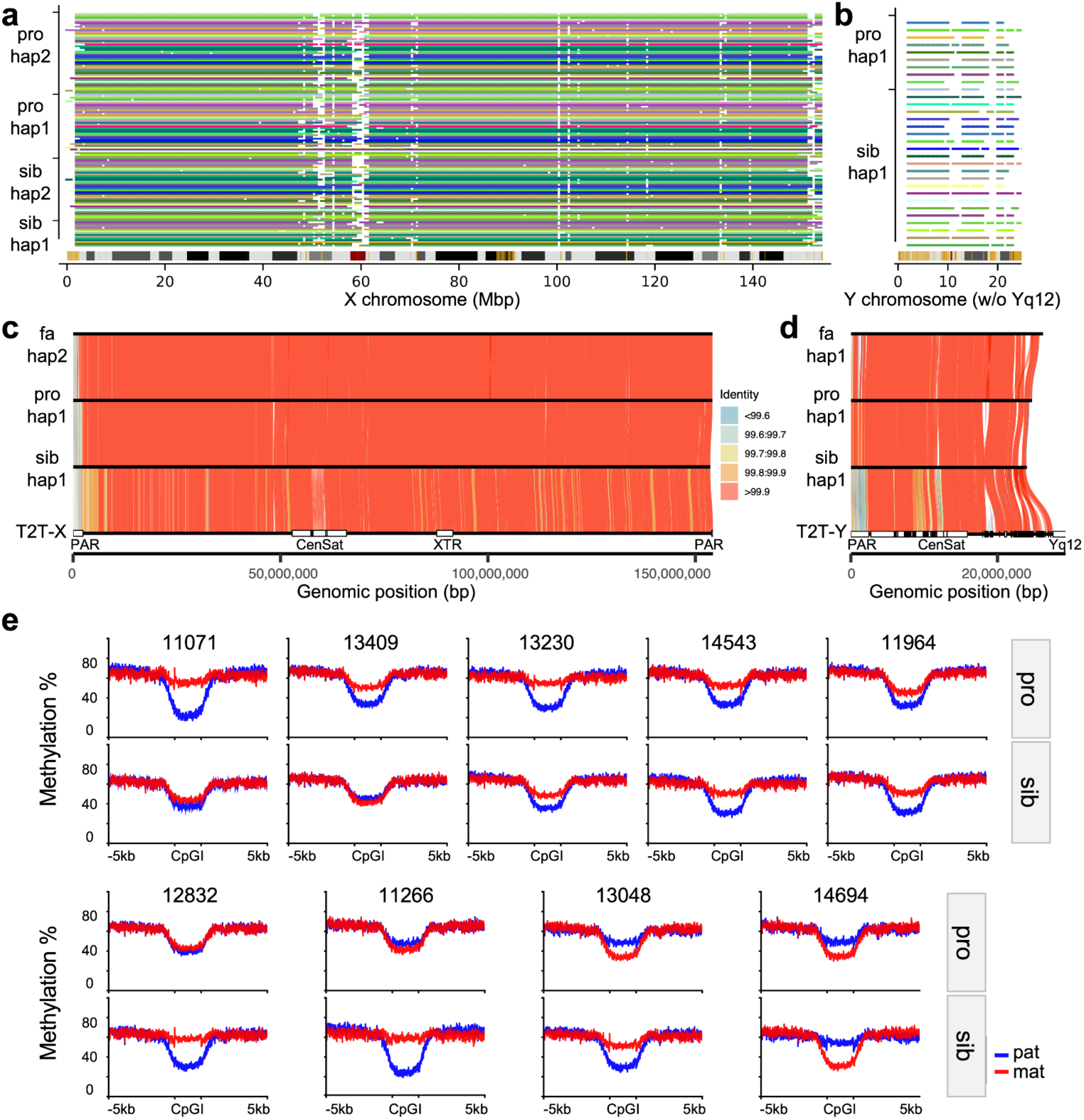
Sex chromosome assembly, transmission and X chromosome inactivation skewing. **a)** Stacked barplot showing X chromosome assembly continuity and mappability relative to the T2T-CHM13v2.0 reference across haplotypes. Each horizontal line represents one haplotype. The assembled contigs in each haplotype traverse the 1 Mbp window of the reference (no more than 3) and have at least ≥95% sequence overlap. Colored segments indicate SDs (yellow), centromeres (red), and gaps (black) on the reference cytogenetic band. **b)** Continuity and mappability of Y chromosome assemblies relative to the T2T-CHM13v2.0 reference (Yq12 heterochromatic region was masked). **c)** Transmitted X assemblies from father to two daughters in 12832 family with sequence identity visualized using gradient colors. **d)** Transmitted Y assemblies from father to sons in 14317 family. Pseudoautosomal regions (PARs), centromeres (Cen) and satellites (Sat), and X-transposed region (XTR) annotations were derived from Rhie et al. (Rhie et al. 2023). **e)** Haplotype-resolved methylation at CpGIs on the X chromosome in nine female-female quads. Mean methylation levels were calculated for each haplotype across 889 CpGIs and their ±5 kbp flanking regions on the X chromosome for 18 female individuals. Red denotes the maternal haplotype, while blue represents the paternal haplotype.

The analysis highlighted three hemizygous tandem repeat (TR) expansion outliers in male probands, each inherited from the mother (**Fig. S8**). These longer TR noncoding variants have only been observed in heterozygous states in females and variants of such lengths have yet to be observed in controls (e.g., *IL1RAPL1* (Interleukin 1 Receptor accessory protein like 1, SFARI score 2 gene). Because LRS data allow CpG methylation to be robustly called (Logsdon et al. 2020), we used the 889 CpG islands across the X chromosomes to assess X chromosome inactivation (XCI) skewing in the blood of female probands and their unaffected sisters. The analysis revealed extreme examples of XCI skewing, including preferential inactivation of the maternally inherited X chromosome (see 11071, **Fig 3e**) potentially consistent with the mother carrying a damaged X chromosome.

### Pathogenic and autism candidate variant discovery

Using the LRS and assembly data, we comprehensively searched for both pathogenic as well as potential candidate mutations missed by SRS analyses (**Methods**). In total, we identified three DNMs classified as pathogenic. Among the 46 idiopathic autism samples, we discovered a *de novo* stop-gain mutation in *SYNGAP1* (**Fig. 4a**), a well-known autism-associated gene encoding a Ras GTPase-activating protein essential for synaptic function and cognitive development (Birtele et al. 2023). This pathogenic DNM in 12237_p1 is clearly supported by both HiFi and ONT reads from the proband but was not reported in three prior SRS analyses of this family (Wilfert et al. 2021; Fu et al. 2022; Satterstrom et al. 2020). An analysis of the Illumina sequence data, however, confirms the presence (**Fig. 4a**) of the variant in a GC-rich region of the genome where a cluster of rare and additional false calls were present, likely resulting in this region being subsequently filtered during QC.

**Figure 4.**
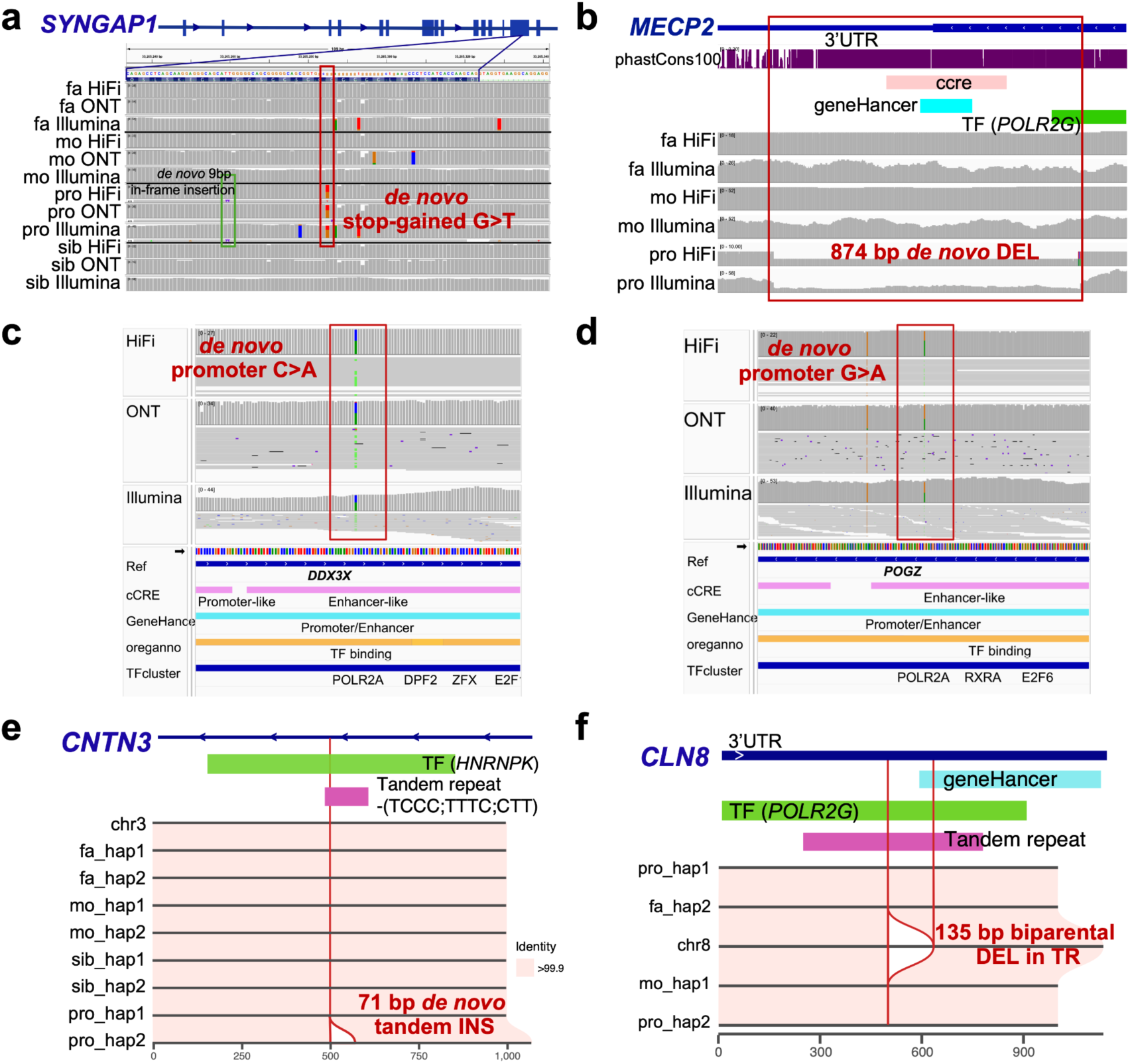
Pathogenic and candidate variants missed by short-read WGS. Long-read sequencing solved cases **a)** (12237_p1) involving a stop-gain *de novo* mutation in *SYNGAP1* and **b)** (HYZ207_p1) involving a *de novo* deletion in the last exon of *MECP2*. **c)** A *de novo* candidate mutation in the promoter of *DDX3X* in 14133_p1. **d)** A *de novo* candidate mutation in the promoter of *POGZ* in 12456_p1. **e)** A 71 bp *de novo* tandem insertion in 11201_p1, predicted to interrupt the HNRNPK TF binding cluster in the intron of *CNTN3*. **f)** A 135 bp homozygous tandem repeat (TR) contraction in the 3’ UTR of *CLN8* in 11616_p1, predicted to disrupt the transcription of *CLN8*.

Two additional pathogenic variants were discovered by LRS among the five autism females with features reminiscent of Rett syndrome. This included an 874 bp *de novo* DEL in *MECP2* in HYZ207_p1, which was largely intronic but effectively disrupts the last exon of the gene (**Fig. 4b**). This pathogenic variant was previously missed in three rounds of clinical testing, including two gene panel sequencing tests through ARUP Laboratories and Quest Diagnostics, and one test of WES through Ambry Genetics. It was confidently identified in our LRS analysis and subsequently validated using all three sequencing platforms. We also identified *de novo* missense mutation within *TBL1XR1* classified by ClinVar as pathogenic (rs1057517933). *TBL1XR1* encodes a transducin (beta)-like 1 X-linked receptor 1–that directly interacts with MECP2. This *de novo TBL1XR1* (D370N) missense variant was previously reported in the same patient (Zaghlula et al. 2018) and two other cases in DECIPHER and have been recently classified as a pathogenic variant (Tillotson and Bird 2020). We also observed a corresponding decrease in methylation at this CpG site within the exon of *TBL1XR1* (**Fig. S9**). As part of this analysis, we also note two DNMs (**Table 1**) called by both SRS and LRS mapping to promoters of genes strongly implicated in neuronal development (*POGZ* and *DDX3X*; **Fig. 4c-d**). *POGZ* encodes a zinc finger protein involved in chromatin remodeling and transcriptional regulation (Stessman et al. 2016), while *DDX3X*, an ATP-dependent RNA helicase, plays a crucial role in RNA metabolism, translation regulation, and neuronal development (Rosa e Silva et al. 2024). Both genes have been implicated in NDDs, including ASD. These two DNMs have never been observed in gnomAD and given their critical location are candidates for functional testing using MPRA to determine if they significantly reduce expression levels.

**Table 1.**
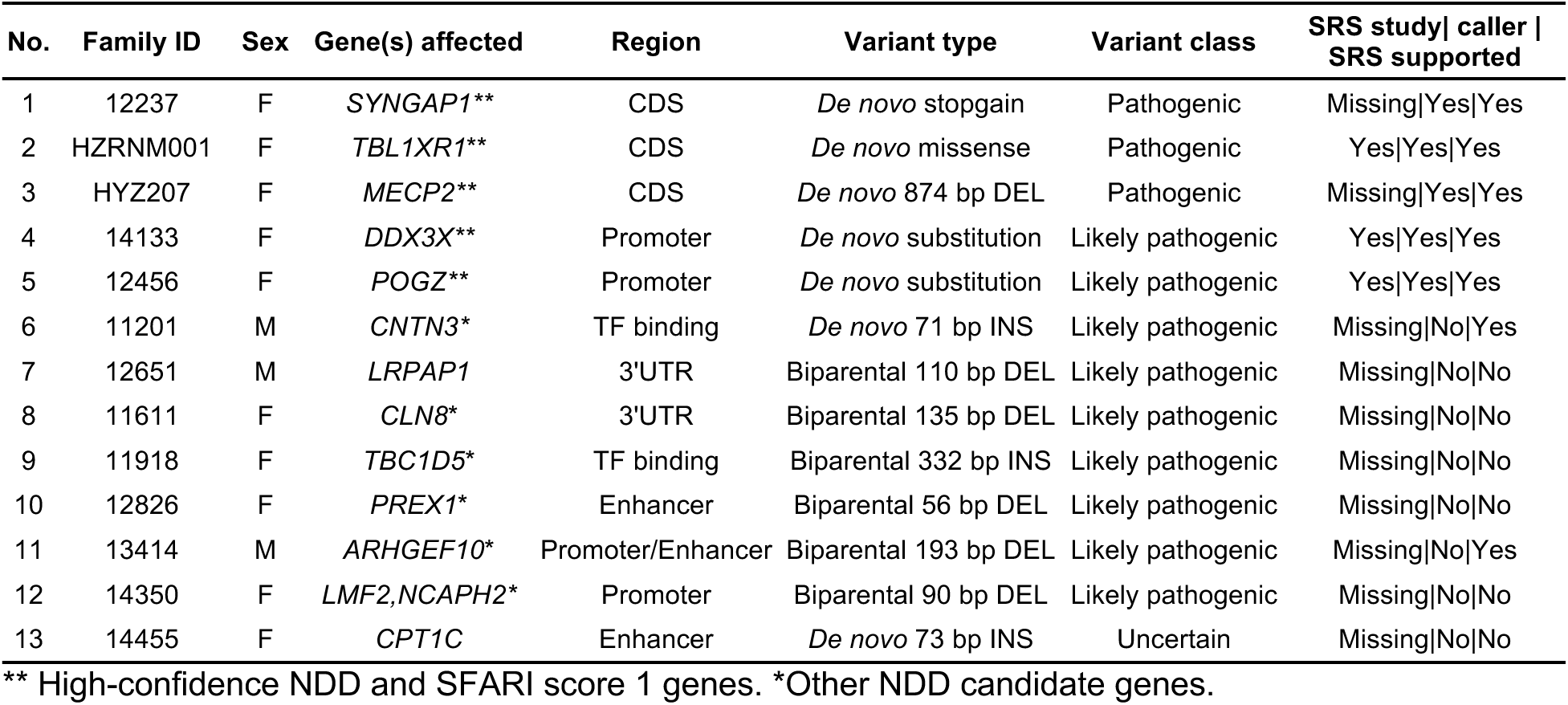
Summary of autism pathogenic and candidate variants.

Among *de novo* SVs, we identified several candidates of potential regulatory consequence (**Table 1**). For example, we identified a 71 bp *de novo* TR INS within the intron of *CNTN3*, a SFARI gene encoding Contactin 3, which mediates cell surface interactions during nervous system development and the outgrowth and guidance of axons and dendrites (**Fig. 4e**). This INS is predicted to disrupt HNRNPK transcription factor (TF) binding sites in 11201_p1. We attempted to recall this SV using SRS-based callers, including Manta (X. Chen et al. 2016), Smoove (https://github.com/brentp/smoove), CNVnator (Abyzov et al. 2011), and Canvas (Roller et al. 2016), and re-genotyped it using Paragraph (S. Chen et al. 2019) based on Illumina alignments. All SRS tools failed to detect this variant, likely due to its mapping within a CT-rich TR. We also identified a 73 bp *de novo* TR INS predicted to interrupt regulatory regions of *CPT1C* and TF binding clusters in 14455_p1. *CPT1C* encodes carnitine palmitoyltransferase 1C, a neuron-specific protein located in the endoplasmic reticulum, and has been associated with spastic paraplegia (Sierra et al. 2008) with an emerging role neuropsychiatric conditions (Kępka et al. 2021). This *de novo* INS was missed by all SRS-based callers likely due to high-GC content (63% within 100 bp).

Finally, we evaluated rare biparental homozygous SVs for potential pathogenicity because of the unique capability of LRS to phase almost all variants (Porubsky, Dashnow, et al. 2025). We identified six candidates SVs inherited biparentally and associated with SFARI risk genes or cortex-specific regulatory regions (**Table 1, Dataset S3**). None of these have been reported as homozygous in over 14,891 genomic controls from gnomAD (Collins et al. 2020) or were identified in pangenome controls. This set includes: a homozygous DEL overlapping an enhancer located in the 3’ untranslated region (UTR) of gene *CLN8* (a SFARI score 2 gene) in proband 11611_p1 (**Fig. 4f**); a homozygous DEL disrupting the enhancer of gene *ARHGEF10* (a SFARI score 2 gene), encoding a Rho guanine nucleotide exchange factor (GEF), in 13414_p1; a homozygous cortex-specific cis-regulatory element DEL in individual 12651_p1 mapping to the 3’ UTR of *LRPAP1*, which encodes LDL receptor-related protein associated protein 1 and has been linked to dementia and late-onset Alzheimer’s disease (Reid and Brown 2023); a homozygous DEL within an intronic enhancer of *PREX1* (a SFARI score 2 gene), in proband 12861_p1; a 332 bp homozygous INS disrupt TF binding in *TBC1D5* (a SFARI score 2 gene) in 11918_p1, and a 500 bp homozygous DEL mapping to the promoter region of both *LMF2* and *NCAPH2* (a SFARI score 3 gene) in individual 14350_p1 (**Table 1**). This promoter region is predicted to harbor regulatory activity based on multiple datasets, including chromatin accessibility and enhancer marks in the developing brain, suggesting potential cis-regulatory effects on the expression of one or both genes.

### Pangenome increased sensitivity for pathogenic SV discovery

To enhance the power of pangenome filtering to effectively exclude common SVs and focus on a high-confidence pool of rare variants, we expanded the control cohort size from 108 to 285 and then to 572 individuals. The 285-control set contains 177 newly LRS and assembled samples from the HPRC (HPRCY2), while the 572-control set includes an additional 287 publicly available 1000 Genomes Project (1KG) samples sequenced using ONT (**Dataset S1, Fig. S3**). Increasing the number of controls, especially samples of African origin, nearly doubles the number of SVs from 271,375 (108 controls) to 445,333 (572 controls) nonredundant SVs. Focusing on rare variants private to a family, the number of rare SVs concomitantly drops to 202 events per sample (**Fig. 5b**). Thus, 99% of common SVs are excluded per individual forming a more tractable and potentially biomedically relevant set of rare variants for downstream interpretation and enrichment analyses. Although still not statistically significant (p=0.23, Mann-Whitney U test), we note that applying 572 controls results in a larger difference with respect to SV burden between probands and unaffected siblings (*Z=1.19*). Of note, only one of our previously 13 proposed candidates (73 bp *de novo* TR INS in *CPT1C*) was excluded and none of our pathogenic variants were excluded (**Table 1, Fig. S10**).

**Figure 5.**
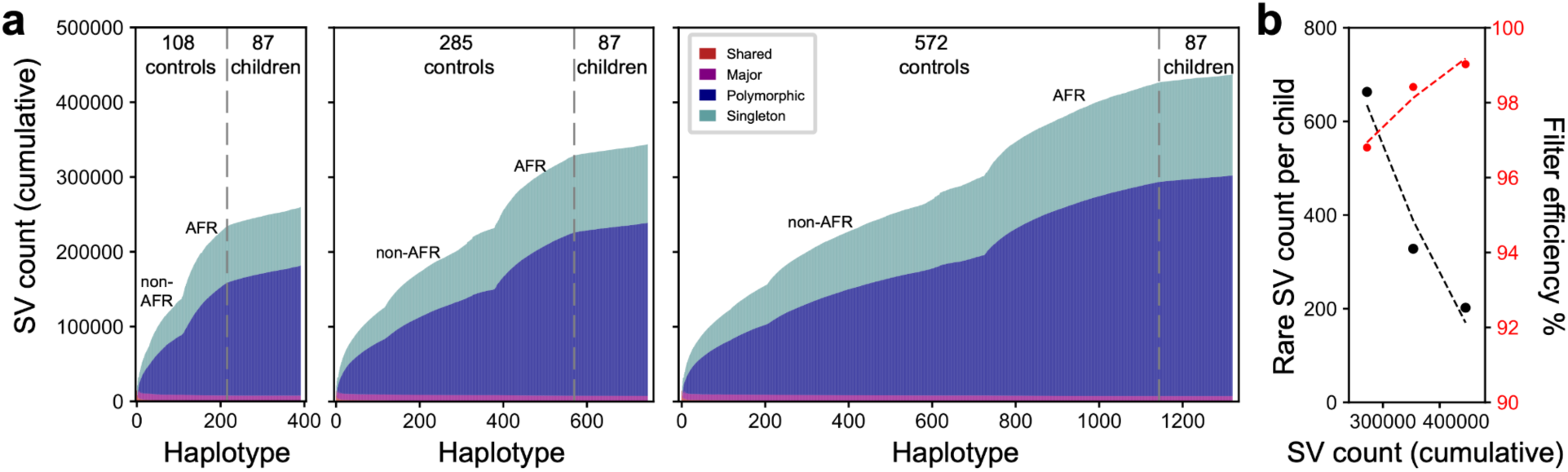
Reduction of the rare SV pool with increasing control samples. **a)** Cumulative discovery curve of SVs identified in different control cohorts of 108, 285 and 572 individuals, compared to 87 children (both affected and unaffected) from autism families. Control samples and discovery curves were computed for both African and non-African controls. **b)** The inclusion of additional population controls refined the rare SV candidate pool, reducing the number of rare SVs (black) from 663 to 202, thus reducing the number of SVs under consideration from 97% to 99% (red).

## DISCUSSION

SRS and microarray studies of autism families have estimated that as much as 30% of autism cases harbor a rare variant of large effect (Yoon et al. 2021; Iossifov et al. 2014). While only approximately half of this burden has been discovered by SRS, it has been hypothesized that missing variants as well as a portion of the missing heritability may be attributed to impactful rare variants mapping in complex regions of the genome that are simply inaccessible or difficult to interpret using SRS approaches (Eichler et al. 2010; Manolio et al. 2009). Targeted LRS studies for missing variants associated with Mendelian disease as well as select families with typically severe NDDs have suggested increases in diagnostic yield ranging from 7.3% to 33% (Hiatt et al. 2024; 2021; Sanchis-Juan et al. 2023; Pauper et al. 2021; Miller et al. 2021). Our analysis of 51 families mostly with daughters affected with autism (often more severely) where we attempted to sequence and assemble the entire euchromatic portion of each genome suggests a much more modest rate of pathogenic variant discovery (5.9%). We consider this yield low given that application of LRS increased genome-wide sensitivity of DNM detection by 20-40% (Noyes et al, 2025) and we purposefully sequence affected females where the probability of discovery of a large effect mutation is expected to be higher (Iossifov et al. 2014).

In the end, we identified three pathogenic variants, including only one daughter previously classified as idiopathic (*SYNGAP1*), while the other two pathogenic mutations (a *de novo* disruptive missense mutation in *TBXLR1* and *de novo* SV affecting the last exon of *MECP2*) arose in daughters suspected of Rett syndrome. A retrospective analysis of whole-genome SRS data confirmed the presence of the variants although in two of the three cases the variants would have been challenging to call without LRS. In addition, we identified nine additional candidate mutations (17.6% of patients) for further functional testing. The majority of these (7/9) were SVs that would have been missed by most standard SRS-based SV callers (**Table 1**). In contrast to DNM, these SV mutations did not map to coding regions but instead were inherited and corresponded to homozygous deletions or insertions within regulatory DNA often for genes associated with autism or neurodevelopment. The LRS data provided unambiguous phasing allowing rare biparental homozygous events to be discovered and characterized. These findings may suggest that some fraction of autism arises as a result of recessive or a contribution of oligogenic mutations (Leblond et al. 2019; Wilfert et al. 2021; Guo et al. 2019). Advanced sequencing techniques such as LRS will be required to reveal the full spectrum of mutations contributing to autism.

Compared to earlier work with large CNVs (Sebat et al. 2007), we do not yet observe a significant increase in SV burden when comparing affected and unaffected siblings (**Fig. 2**). This lack of statistical significance is likely due to the limited sample size of this study. Notwithstanding, there are some interesting trends. For example, we note a slight excess of deletions on the X chromosome among affected daughters when compared to their unaffected sisters. Indeed, our ability to sequence and assemble ∼96% of the X chromosome, as well as most of the euchromatic portion of the Y chromosome (**Fig. 3**), will be critical for evaluating the contribution of sex chromosomes to autism sex bias. Sex chromosomes are routinely excluded from SRS-WGS studies because of ploidy issues and challenges with repetitive regions (Fu et al. 2022; Satterstrom et al. 2020; Trost et al. 2020). LRS and assembly largely overcome these limitations. Moreover, the ability to concurrently assay methylation status of CpG islands genome-wide and readily distinguish X chromosome skewing patterns (**Fig. 3**) will also advance the discovery of epigenetic mutations as well as potentially damaged X chromosomes as more and more autism genomes are sequenced.

Perhaps, most importantly has been the ability to leverage pangenomes (Liao et al. 2023) to restrict the focus of SV discovery to variants that are private to autism families. Unlike SNVs mapping to coding sequencing, databases such as gnomAD (Collins et al. 2020; S. Chen et al. 2024; Karczewski et al. 2020) are largely incomplete for variants, especially SVs, mapping to more complex regions of the genome. A typical human genome harbors over 25,000 variants while whole-genome SRS has been shown to reliably report only 11,000 such variants (Chaisson et al. 2019). In this study, we used more completely sequenced and assembled genomes from public initiatives such as HGSVC, HPRC and 1KG ONT (Liao et al. 2023; Logsdon, Ebert, et al. 2024; Ebert et al. 2021; Gustafson et al. 2024) as controls to filter out more common SVs. Using 572 LRS control genomes identifies 445,333 nonredundant SVs in total. Under a model of ultra-rare SVs contributing to disease, we, as a result, essentially exclude 99% of the more common variants allowing us to focus on 202 private or *de novo* SV variants per child. We note that differential with respect to SV burden increases between proband and unaffected sibling although does not yet reach statistical significance. If we further restrict this analysis to SVs corresponding to regions of the genome under functional constraint (with Gnocchi ≥ 4 (S. Chen et al. 2024) and overlap with predicted promoters), this set would further reduce to one or two SVs per genome. As the human pangenome continues to grow and more complete genetic information emerges, the potential to discover variants of pathogenic significance will increase.

## MATERIALS AND METHODS

### Sample selection and previous sequence characterization

Illumina WGS was previously applied to the blood DNA of the 174 individuals corresponding to 46 families with idiopathic autism from the SSC and SAGE. Potential pathogenic variants were screened from SRS based on the studies by Wilfert et al. (Wilfert et al. 2021) and Fu et al. (Fu et al. 2022), including (1) SNVs: de novo likely gene-disruptive (LGD), de novo missense and rare inherited LGD variants in NDD-related genes or genes with pLI scores ≥ 0.9; (2) CNVs in the morbidity map (Coe et al. 2019) or span of a gene with pLI score ≥ 0.9. No known genetic cause was identified in the 46 probands from SRS data (Fu et al. 2022; Wilfert et al. 2021; Coe et al. 2019; Turner et al. 2017). SNVs were identified by GATK (Poplin, Ruano-Rubio, et al. 2018), FreeBayes (Garrison and Marth 2012), Platypus (Rimmer et al. 2014) and Strelka2 (Illumina). CNVs were called by GATK-gCNV (Poplin, Ruano-Rubio, et al. 2018), GATK-SV (Collins et al. 2020), WhamG (Kronenberg et al. 2015), Lumpy (Layer et al. 2014), Delly2 (Rausch et al. 2012), GenomeSTRiP (Handsaker et al. 2015), dCGH (Sudmant, Mallick, et al. 2015) and CNVnator (Abyzov et al. 2011). In addition, the 46 probands selected in this study do not exhibit exceptional polygenic risk scores among the 13,989 individuals examined (Wilfert et al. 2021) and have known IQs ranging from 13 to 91. Similarly, five girls diagnosed with Rett-like syndrome from Baylor College of Medicine had no causal variants identified in *MECP2* by prior gene panel testing or WES (**Dataset S1**). The selected families were predominantly those with female probands due to the interest in discovering X chromosome variants and the large-effect variants are more likely to be discovered. As shown in **Fig. 1a**, 17 quads with sex-matched offspring (12 female-female and 5 male-male quads), 19 quads with sex-mismatched offspring (16 female-male and 3 male-female quads) and 17 trios (15 female and 2 male trios) were included in this study. Briefly, the 51 probands (41 females and 10 males) were selected to represent cases that pose difficulties in pinpointing the cause of ASD using Illumina SRS data.

### LRS data generation and QC

We generated PacBio HiFi and ONT sequencing data at the University of Washington (UW) for 189 individuals. Illumina WGS for Rett-like trios was generated from blood DNA using the TruSeq library kit and sequenced on a NovaSeq with paired-end 150 bp reads at the Northwest Genomics Center. For the five Rett-like trios, DNA was extracted from blood using the Monarch HMW DNA Extraction Kit for Cells & Blood from NEB (T3050L) (n=6) or the Qiagen Puregene Blood Core Kit (158023) (n=9), following manufacturer’s specifications. The whole-blood DNA from the SSC was extracted previously as part of that biobank. The cell line DNA was extracted from lymphoblastoid cell lines with either a modified Gentra Puregene (Qiagen) protocol when used for ONT sequencing or with the Monarch HMW kit (NEB T3050L) when used for PacBio sequencing. ONT libraries were constructed using the Ligation Sequencing Kit (ONT, LSK110 and LSK114) with modifications to the manufacturer’s protocol. The library was loaded onto a primed R9.4.1 or R10.4.1 flow cell (FLO-PRO002 or FLO-PRO114M) for sequencing on the PromethION, with two nuclease washes and reloads after 24 and 48 hours of sequencing.

PacBio HiFi data from family 14455 (n=4) were published in Noyes et al. 2022 (Noyes et al. 2022). Data from three families (n=11) were graciously generated by PacBio. Remaining individuals’ HiFi data were generated from blood or cell line HMW DNA according to the manufacturer’s recommendations. At all steps, quantification was performed with Qubit dsDNA HS (Thermo Fisher Scientific, Q32854) measured on DS-11 FX (Denovix) with the size distribution checked using FEMTO Pulse (Agilent, M5330AA and FP-1002-0275.) The samples’ incoming size distribution determined shearing conditions, either no shear (n=12), or sheared with the Megaruptor 3 (Hologic Diagenode, B06010003 & E07010003) system using one (n=36) or two (n=130) sequential runs to target a peak size of ∼20 kbp. After shearing, the DNA were used to generate PacBio HiFi libraries using the Express Template Prep Kit v2 (n=12, PacBio, 100-938-900) or SMRTbell prep kit 3.0 (n=166, PacBio, 102-182-700). Size selection was performed with Pippin HT using a high-pass cut-off between 9-17 kbp based on shear size (Sage Science, HTP0001 and HPE7510). Libraries were sequenced either on the Sequel II platform on SMRT Cells 8M (PacBio, 101-389-001) using Sequel II sequencing chemistry 2.0 (n=16, PacBio, 101-842-900), 2.2 (n=4, PacBio, 102-089-000), or 3.2 (n=32, PacBio,102-333-300) with 2 h pre-extension and 30 h movies on SMRT Link v. 9-11.1, or on the Revio platform on Revio SMRT Cells (PacBio, 102-202-200) and Revio polymerase kit v1 (n=126, PacBio, 102-817-600) with 2 h pre-extension and 24 or 30 h movies on SMRT Link v.12.0-13.1.

To ensure the use of high-quality reads for constructing robust assemblies, we first filtered out nonhuman contamination reads from both HiFi and ONT data. We employed highly accurate Illumina reads and utilized yak (https://github.com/lh3/yak.git, commit f389bad) to calculate the quality value (QV) for each read, and a z-test was conducted on the resulting QV values. Reads with a z-score less than -2, indicating a potential risk of contamination, were compared against the Kraken2 (v2.1.3, (Wood et al. 2019)) database, and those identified as nonhuman in origin were excluded. We masked the Y chromosome from GRCh38 to generate the GRCh38noY reference genome. HiFi reads were aligned to GRCh38 for males and to GRCh38noY for females using pbmm2 (v1.13.1, https://github. com/PacificBiosciences/pbmm2). To ensure the family pedigree, the relatedness between sample pairs and ancestry prediction were conducted using Somalier (v0.2.19, (Pedersen et al. 2020)) based on the alignment (**Dataset S1**). ONT reads were aligned to the reference genome using minimap2 (v2.28.0, (Li 2021)), and the family pedigree was confirmed with VerifyBamID (v2.0.1, (Zhang et al. 2020)). In addition, ntsm (v1.2.1, (Chu et al. 2024)) was applied to each HiFi and ONT fastq for sample swap detection.

### Phased genome assembly construction

The HiFi assembly was constructed by hifiasm (v0.16.1, (Cheng et al. 2024)). Parental short reads were processed with yak (v0.1, https://github.com/lh3/yak.git) and then hifiasm trio-binning mode was used for phasing children samples, while the parental assemblies were partially phased by default. The sex chromosome contigs from father samples were aligned to the T2T-CHM13v2.0 reference to reassign the Y chromosome contigs to hap1 (or paternal haplotype) and X chromosome contigs to hap2 (or maternal haplotype). Assembly QVs were evaluated by merqury (v1.3, (Rhie et al. 2020)) with k-mers from Illumina data (meryl v1.4, (Rhie et al. 2020)); next, the completeness of phased assemblies relative to the reference and contig N50 values were calculated (**Dataset S1**).

### Variant discovery

SNVs and small indels in 73 children, with available ONT data, were recalled with DeepVariant (v1.4.0, (Poplin, Chang, et al. 2018)) and GATK (v4.3.0.0, (Poplin, Chang, et al. 2018)) based on the HiFi alignments to the T2T-CHM13v2.0 reference. DNMs including both *de novo* and postzygotic mutations were further validated by ONT and/or Illumina reads using the method described in Noyes et al. 2025. We annotated DNMs using the Ensembl Variant Effect Predictor (VEP, v110.1, (McLaren et al. 2016)) and referred to the predicted impact and scores from dbNSFP (v4.8a, (Liu et al. 2020), CADD score (v1.3, (Kircher et al. 2014), and gnomAD genome allele frequency (v4.1.0, (S. Chen et al. 2024) by lifting coordinates over to both GRCh38 and GRCh37 using UCSC LiftOver (Hinrichs et al. 2006) (**Dataset S3**). SVs were called using the phased assembly variant caller (PAV, v2.3.4, (Ebert et al. 2021; Audano et al. 2019)) by aligning the assembled genomes to a reference (GRCh38 or GRCh38noY). The alignment-based SVs were called by PacBio SV calling and analysis tool (PBSV, v2.9.0, https://github.com/PacificBiosciences/pbsv) and Sniffles (v2.2, Smolka et al. 2024). SVs from the control population were additionally called by Delly (v1.2.6, Rausch et al. 2012), Sawfish (v0.12.4, Saunders et al. 2024), and cuteSV (v2.1.0, Jiang et al. 2020). The ratio of insertions (INS) to deletions (DEL), the ratio of heterozygous (HET) to homozygous (HOM) variants, and the size distribution of SVs were evaluated for each VCF file.

### SV merging and filtering strategy

The merging strategy involved two major steps. Briefly,

1. Callerset validation. SVs detected in each sample from different SV callers were first normalized, sorted, and merged with BCFtools (v1.20, (Danecek et al, 2021)) on the basis of PAV callset, and then collapsed by Truvari (v4.3.1, (English et al. 2022)). SVs located within known genomic gaps, telomeric regions, centromeres, and PARs on GRCh38 from UCSC Genome Browser tracks were excluded. We used the following command line:

bcftools merge --thread {threads} --merge none --force-samples -O z -o {output.vcf.gz} {input.vcf1.gz} {input.vcf2.gz} {input.vcf3.gz}

truvari collapse -i {input.vcf.gz} -c {output.removed.vcf.gz} --sizemin 0 --sizemax 1000000 -k maxqual --gt het --intra --pctseq 0.90 --pctsize 0.90 --refdist 500 | bcftools sort --max-mem 8G - O z -o {output.collapsed.vcf.gz}

2. Inter-sample merge. We extracted SVs supported by PAV and at least one of the alignment-based callers for each individual and then merged SVs from both controls and autism families using a list of VCFs with Truvari. In this study, we have applied three control sets consisting of 108, 285, and 572 individuals.

bcftools merge --threads {threads} --merge none --force-samples --file-list {input.vcflist} -O z | bcftools norm --threads 15 --do-not-normalize --multiallelics -any --output-type z -o {output.mergevcf.gz}

truvari collapse --input {input.mergevcf.gz} --collapsed-output {output.removed_vcf.gz} --sizemin 0 --sizemax 1000000 --pctseq 0.90 --pctsize 0.90 --keep common --gt all | bcftools sort --max-mem {resources}G --output-type z > {output.collapsed_vcf.gz}

3. Rare SV pool discovery. We developed a custom script to extract six categories of rare SVs as described in the main text (https://github.com/EichlerLab/asap).

### Transmission curation of rare SVs

For autosomal SVs, we retained heterozygous and homozygous SVs present only in the children but not in the controls. For SVs on the sex chromosomes, we performed sex-matched comparisons and filtered SVs seen in controls with the same sex. Parental genotypes corresponding to each rare SV in the children are provided in

### Dataset S2

Variants that followed Mendelian inheritance patterns were designated as high-confidence SVs. The remaining SVs were subjected to transmission curation using the following toolchain:

1. Initial caller support using Truvari: To minimize the loss of inheritance information in parents lacking SV caller support, we collapsed SVs from parents with the child’s SVs using truvari bench,

truvari bench -c {fa,mo}.vcf.gz -b child.vcf --pctsize 0.9 --pctseq 0.9 -o {fa_child,mo_child}

2. Callable region evaluation using BoostSV: To ensure the SVs fall within confidently callable regions across samples in a single family, we developed a tool, BoostSV (https://github.com/jiadong324/BoostSV), leveraging a machine-learning approach trained on control samples (Porubsky, Dashnow, et al. 2025). This tool evaluates read support, mapping quality, and data quality metrics from alignments surrounding the target SVs in each parent. A quality threshold of 0.5 was applied to obtain the transmission.

3. Genotyping support using kanpig: We applied the k-mer-based genotyper kanpig (v0.3.1, (English et al. 2025)) to parental HiFi alignments for each SV to assess allele presence and genotype consistency.

4. Rare TR expansions/contractions and multiple sequence alignment (MSA): SVs overlapped with TR catalogs derived from the four-generation control family in Porubsky et al. (Porubsky, Dashnow, et al. 2025) were genotyped by TRGT (v1.4.1-e655c85, (Dolzhenko et al. 2024)) using,

trgt genotype --genome {input.ref} --repeats {input.bed} --reads {input.bam} -t {threads} --output-prefix {wildcards.sample}/trgt --karyotype {XX/XY}

Genotype (GT) and allele length (AL) information were extracted from the TRGT output across all individuals of the ASD family to detect transmission. For complex TR motif structures, we validated the SVs in the target sequence from assemblies using the MSA approach,

\# Align assemblies to the reference using minimap2 (v2.28.0, (Li 2021)):

minimap2 -c -t {threads} -K {resources.mem}G --cs -x asm20 -m 10000 -z 10000,50 -r 50000 -- end-bonus=100 -O 5,56 -E 4,1 -B 5 --secondary=no --eqx -Y {input.ref} {input.asm} > {output.paf}

\# Liftover target sequence coordinates onto query sequence using rustybam (v0.1.33, https://github.com/mrvollger/rustybam) and extract the target sequence using samtools (v1.16.1, (Danecek et al. 2021)):

rustybam liftover --bed {input.bed} {input.paf} > {output.liftover.paf} samtools faidx {input.asm} {liftover.paf.query_region} > {output.fa}

#. MSA were performed using MAFFT (v7.525, (Katoh et al. 2002)) and visualized in Jalview (v2.11.4.1, (Waterhouse et al. 2009)):

mafft --adjustdirection --thread {threads} --auto --reorder {input.combined.fa} > {output.msa.fa}

5. Read-based support validation using subseq: To further assess SV transmission, subseq (https://github.com/EichlerLab/subseq, (Ebert et al. 2021)), was used to quantify read support for each SV in parental genomes. A dynamic window size was determined based on the SV size, and the number of reads traversing the window counted. SVs with ≥3 supporting reads were considered moderate-confidence transmissions, while those with a single supporting read were marked as low-confidence.

6. Manual inspection using IGV: For SVs lacking sufficient support in Steps 1-4, we conducted visual inspection of supporting reads in the Integrated Genomics Viewer (IGV, v2.16.0, (Robinson et al. 2017)). Evidence from HiFi alignments, HiFi assemblies, Illumina alignments, and, when available, ONT alignments (152 samples) was reviewed to further assess inheritance status and assign low-confidence transmissions.

### SV annotation

A customized script was implemented to annotate previously published NDD candidate genes and regulatory elements (REG), as well as integrate annotations from AnnotSV (v3.4). In terms of REG, we integrated published datasets from UCSC Genome Browser tracks (Perez et al. 2025), including candidate cis-regulatory elements (ENCODE Regulation, ENCODE cCREs, ORegAnno, GeneHancer), ENCODE histone marks (H3K27Ac, H3K4Me1, H3K4Me3), and ENCODE TF Clusters. Additionally, we incorporated epigenomic profiles from the cerebral cortex (brainREG), particularly a cis-regulatory element map generated from 27 male and 21 female prenatal human cortex samples by ATAC-seq and consensus maps for CTCF, H3K27ac, H3K27me3, and H3K4me3 generated from six male and five female prenatal human cortex samples by CUT&Tag. The brainREG annotation resulted in a 45% increase in the total number of regulatory SVs. For NDD candidate genes, we primarily focused on those previously reported by SFARI Gene, Fu et al., Zhou et al., Satterstrom et al. and Wang et al. (Fu et al. 2022; Wang et al. 2022; Satterstrom et al. 2020). Additional UCSC noncoding RNA (tRNA, snRNA, lincRNA, sno_miRNA), repetitive regions (UCSC SegDup, UCSC RepeatMasker, UCSC Simple Repeats, TRs (Porubsky, Dashnow, et al. 2025)), noncoding constraint Gnocchi score (S. Chen et al. 2024) and CADD-SV score (v1.1.2, (Kleinert and Kircher 2022)) were annotated.

Potential pathogenic variants were confirmed with gnomAD allele frequency and the 572-control dataset. SVs and CNVs from Illumina WGS data for the selected samples were recalled by Manta (v.1.5.0, (X. Chen et al. 2016)), Smoove (v0.2.5, https://github.com/brentp/smoove), CNVnator (v0.3.3, (Abyzov et al. 2011)), Canvas (v1.40.0.1613+master, (Roller et al. 2016)) and genotyped using Paragragh (v2.4, (S. Chen et al. 2019)) to evaluate SRS detection.

### Sex chromosome assemblies and transmission

We partitioned the sex chromosomes (T2T-CHM13v2.0, excluding the PAR, centromere, and extremely repetitive Yq12 heterochromatin regions) into 1 Mbp windows and identified those covered by contigs that aligned to ≥95% of the window sequence with no more than three overlapping contigs. The coverage percentage was calculated as the number of qualified windows divided by the total number of windows, representing coverage relative to the reference sex chromosomes (Fig. 3a-b, Fig. S7). We designed a pipeline to assemble contiguous X and Y chromosomes and validate transmission patterns within families (https://github.com/projectoriented/contiguous-X). The pipeline consists of two main steps: (1) scaffolding contigs with at least 50% of their sequence aligning to chromosome X (minimap2 (Li 2021)) via RagTag (v2.1.0, (Alonge et al. 2022)), and (2) visualizing alignments across haplotypes within the family using the SVbyEye R package (Porubsky, Guitart, et al. 2025).

### Methylation analysis

We developed a pipeline to extract phased methylation signals from ONT alignments (https://github.com/projectoriented/continuous-methylation?tab=readme-ov-file). Briefly, when parental Illumina reads were available, ONT reads from the offspring were phased using the Canu (v2.1.1, (Koren et al. 2018)) trio-binning method. The phased ONT reads were then aligned to the GRCh38 reference genome using minimap2 (v2.24.0, (Li 2021)) and haplotagged accordingly. For individuals (e.g., parents) without available parental Illumina data, SVs were identified using Sniffles (v2.2, (Smolka et al. 2024)) and small variants were called with Clair3 (v1.0.2, (Zheng et al. 2022)), followed by haplotagging with LongPhase (v1.7.2, (Lin et al. 2022)). Methylation tags from unmapped BAM files were linked to the phased alignments using methylink (v0.6.0, https://github.com/projectoriented/methylink). Finally, Modkit (v0.3.1, https://github.com/nanoporetech/modkit) was used to generate methylation calls in BED format via pileup function. We analyzed mean methylation differences between the two X chromosome haplotypes across 889 CpG islands, including ±5 kb flanking regions. We then used deepTools (v3.5.5, https://github.com/deeptools/deepTools) to compute the methylation matrix and generate the plots.

## Data Availability

The data used for analysis are available in the SFARI Base under the accession number SFARI_DS714840 and in the NDA Collection ID 3780.

https://nda.nih.gov/

https://base.sfari.org

## Ethics

Ethical approval for this study was granted by the University of Washington IRB Committee B, under STUDY ID: STUDY00000383.

## Code availability

All pipelines/scripts/custom code/software generated as part of this analysis are available in GitHub at https://github.com/EichlerLab/asap. Any additional information required to reanalyze the data reported in this work paper is available from the lead contact upon request.

## Acknowledgments

We thank all of the individuals who participated in this research. We also thank all contributing investigators to the consortia datasets used here from SSC, SAGE and Baylor College of Medicine, and the families who participated in this research, without whose contributions genetic studies would be impossible. We thank Tonia Brown for assistance in editing this manuscript. We thank Tom Mokveld (PacBio) for helpful discussions. This work was supported, in part, by the US National Institutes of Health (NIH R01MH101221 to E.E.E.; R01NS057819 to H.Y.Z.; 1F32HD116501-01 to R.M-S.; and DP5OD033357 to D.E.M.) and the Simons Foundation (SFARI #810018 to E.E.E., H.Y.Z., T.J.N., and A.C.). E.E.E. and H.Y.Z. are investigators of the Howard Hughes Medical Institute. This work was also supported, in part, by the National Natural Science Foundation of China (82201314 and 82471194) to T.W. This article is subject to HHMI’s Open Access to Publications policy. HHMI lab heads have previously granted a nonexclusive CC BY 4.0 license to the public and a sublicensable license to HHMI in their research articles. Pursuant to those licenses, the author-accepted manuscript of this article can be made freely available under a CC BY 4.0 license immediately upon publication.

## Consortia

Human Pangenome Reference Consortium (HPRC) (Supplemental Note)

## Author contributions

Conceptualization: E.E.E., Y.S.

Sample Collection: K.Ho., R.A.P.P., D.P., B.S.

Data Generation: K.M.M., K.Ho., G.H.G., J.K.

Data QC: Y.S., Y.K., I.W., N.K.

Formal Analysis: Y.S., J.L., M.D.N., I.W., N.K.

Visualization: Y.S., I.W.

Methodology: Y.S., J.L., W.T.H., M.W.

Resources: K.He., D.K., J.A.G., D.E.M., HPRC

Software: Y.K., I.W., N.K., J.W.

Validation: J.L., I.W., T.W., R.M.-S., F.C.

Writing – Original Draft: Y.S.

Writing – Review & Editing: Y.S., E.E.E., H.Y.Z., J.L., M.D.N., Y.K., I.W., N.K., W.T.H., M.W., J.W., K.Ho, K.M.M., G.H.G., J.K., T.W., K.He., D.K., R.A.P.P., R.M-S., F.C., D.P., B.S., J.A.G., D.E.M., T.J.N., A.C., H.B-R., HPRC

Supervision: E.E.E., H.Y.Z., A.C., T.J.N.

Funding Acquisition: E.E.E., H.Y.Z., D.E.M., R.M-S.

## References

Abyzov, Alexej, Alexander E. Urban, Michael Snyder, and Mark Gerstein. 2011. “CNVnator: An Approach to Discover, Genotype, and Characterize Typical and Atypical CNVs from Family and Population Genome Sequencing.” Genome Research 21 (6): 974–84. 10.1101/gr.114876.110.

Alonge, Michael, Ludivine Lebeigle, Melanie Kirsche, et al. 2022. “Automated Assembly Scaffolding Using RagTag Elevates a New Tomato System for High-Throughput Genome Editing.” Genome Biology 23 (1): 258. 10.1186/s13059-022-02823-7.

Audano, Peter A., Arvis Sulovari, Tina A. Graves-Lindsay, et al. 2019. “Characterizing the Major Structural Variant Alleles of the Human Genome.” Cell 176 (3): 663–675.e19. 10.1016/j.cell.2018.12.019.

Birtele, Marcella, Ashley Del Dosso, Tiantian Xu, et al. 2023. “Non-Synaptic Function of the Autism Spectrum Disorder-Associated Gene SYNGAP1 in Cortical Neurogenesis.” Nature Neuroscience 26 (12): 2090–103. 10.1038/s41593-023-01477-3.

Carvalho, Claudia M. B., and James R. Lupski. 2016. “Mechanisms Underlying Structural Variant Formation in Genomic Disorders.” Nature Reviews Genetics 17 (4): 224–38. 10.1038/nrg.2015.25.

Chaisson, Mark J. P., Ashley D. Sanders, Xuefang Zhao, et al. 2019. “Multi-Platform Discovery of Haplotype-Resolved Structural Variation in Human Genomes.” Nature Communications 10 (1): 1784. 10.1038/s41467-018-08148-z.

Chen, Sai, Peter Krusche, Egor Dolzhenko, et al. 2019. “Paragraph: A Graph-Based Structural Variant Genotyper for Short-Read Sequence Data.” Genome Biology 20 (1): 291. 10.1186/s13059-019-1909-7.

Chen, Siwei, Laurent C. Francioli, Julia K. Goodrich, et al. 2024. “A Genomic Mutational Constraint Map Using Variation in 76,156 Human Genomes.” Nature 625 (7993): 92–100. 10.1038/s41586-023-06045-0.

Chen, Xiaoyu, Ole Schulz-Trieglaff, Richard Shaw, et al. 2016. “Manta: Rapid Detection of Structural Variants and Indels for Germline and Cancer Sequencing Applications.” Bioinformatics 32 (8): 1220–22. 10.1093/bioinformatics/btv710.

Cheng, Haoyu, Mobin Asri, Julian Lucas, Sergey Koren, and Heng Li. 2024. “Scalable Telomere-to-Telomere Assembly for Diploid and Polyploid Genomes with Double Graph.” Nature Methods 21 (6): 967–70. 10.1038/s41592-024-02269-8.

Cheng, Haoyu, Gregory T. Concepcion, Xiaowen Feng, Haowen Zhang, and Heng Li. 2021. “Haplotype-Resolved de Novo Assembly Using Phased Assembly Graphs with Hifiasm.” Nature Methods 18 (2): 170–75. 10.1038/s41592-020-01056-5.

Chu, Justin, Jiazhen Rong, Xiaowen Feng, and Heng Li. 2024. “Ntsm: An Alignment-Free, Ultra-Low-Coverage, Sequencing Technology Agnostic, Intraspecies Sample Comparison Tool for Sample Swap Detection.” GigaScience 13 (January): giae024. 10.1093/gigascience/giae024.

Coe, Bradley P., Holly A. F. Stessman, Arvis Sulovari, et al. 2019. “Neurodevelopmental Disease Genes Implicated by de Novo Mutation and Copy Number Variation Morbidity.” Nature Genetics 51 (1): 106–16. 10.1038/s41588-018-0288-4.

Collins, Ryan L., Harrison Brand, Konrad J. Karczewski, et al. 2020. “A Structural Variation Reference for Medical and Population Genetics.” Nature 581 (7809): 444–51. 10.1038/s41586-020-2287-8.

Collins, Ryan L., and Michael E. Talkowski. 2025. “Diversity and Consequences of Structural Variation in the Human Genome.” Nature Reviews Genetics, January 21, 1–20. 10.1038/s41576-024-00808-9.

Danecek, Petr, James K Bonfield, Jennifer Liddle, et al. 2021. “Twelve Years of SAMtools and BCFtools.” GigaScience 10 (2): giab008. 10.1093/gigascience/giab008.

Dolzhenko, Egor, Adam English, Harriet Dashnow, et al. 2024. “Characterization and Visualization of Tandem Repeats at Genome Scale.” Nature Biotechnology 42 (10): 1606–14. 10.1038/s41587-023-02057-3.

Ebert, Peter, Peter A. Audano, Qihui Zhu, et al. 2021. “Haplotype-Resolved Diverse Human Genomes and Integrated Analysis of Structural Variation.” Science 372 (6537): eabf7117. 10.1126/science.abf7117.

Eichler, Evan E., Jonathan Flint, Greg Gibson, et al. 2010. “Missing Heritability and Strategies for Finding the Underlying Causes of Complex Disease.” Nature Reviews Genetics 11 (6): 446–50. 10.1038/nrg2809.

English, Adam C., Fabio Cunial, Ginger A. Metcalf, Richard A. Gibbs, and Fritz J. Sedlazeck. 2025. “K-Mer Analysis of Long-Read Alignment Pileups for Structural Variant Genotyping.” Nature Communications 16 (1): 3218. 10.1038/s41467-025-58577-w.

English, Adam C., Vipin K. Menon, Richard A. Gibbs, Ginger A. Metcalf, and Fritz J. Sedlazeck. 2022. “Truvari: Refined Structural Variant Comparison Preserves Allelic Diversity.” Genome Biology 23 (1): 271. 10.1186/s13059-022-02840-6.

Fu, Jack M., F. Kyle Satterstrom, Minshi Peng, et al. 2022. “Rare Coding Variation Provides Insight into the Genetic Architecture and Phenotypic Context of Autism.” Nature Genetics 54 (9): 1320–31. 10.1038/s41588-022-01104-0.

Garrison, Erik, and Gabor Marth. 2012. “Haplotype-Based Variant Detection from Short-Read Sequencing.” arXiv:1207.3907. Preprint, arXiv, July 20. 10.48550/arXiv.1207.3907.

Guo, Hui, Michael H. Duyzend, Bradley P. Coe, et al. 2019. “Genome Sequencing Identifies Multiple Deleterious Variants in Autism Patients with More Severe Phenotypes.” Genetics in Medicine 21 (7): 1611–20. 10.1038/s41436-018-0380-2.

Gustafson, Jonas A., Sophia B. Gibson, Nikhita Damaraju, et al. 2024. “Nanopore Sequencing of 1000 Genomes Project Samples to Build a Comprehensive Catalog of Human Genetic Variation.” Preprint, medRxiv, March 7. 10.1101/2024.03.05.24303792.

Handsaker, Robert E., Vanessa Van Doren, Jennifer R. Berman, et al. 2015. “Large Multiallelic Copy Number Variations in Humans.” Nature Genetics 47 (3): 296–303. 10.1038/ng.3200.

Hiatt, Susan M., James M. J. Lawlor, Lori H. Handley, et al. 2021. “Long-Read Genome Sequencing for the Molecular Diagnosis of Neurodevelopmental Disorders.” Human Genetics and Genomics Advances 2 (2): 100023. 10.1016/j.xhgg.2021.100023.

Hiatt, Susan M., James M. J. Lawlor, Lori H. Handley, et al. 2024. “Long-Read Genome Sequencing and Variant Reanalysis Increase Diagnostic Yield in Neurodevelopmental Disorders.” Genome Research 34 (11): 1747–62. 10.1101/gr.279227.124.

Hinrichs, A. S., D. Karolchik, R. Baertsch, et al. 2006. “The UCSC Genome Browser Database: Update 2006.” Nucleic Acids Research 34 (suppl_1): D590–98. 10.1093/nar/gkj144.

Iossifov, Ivan, Brian J. O’Roak, Stephan J. Sanders, et al. 2014. “The Contribution of de Novo Coding Mutations to Autism Spectrum Disorder.” Nature 515 (7526): 216–21. 10.1038/nature13908.

Karczewski, Konrad J., Laurent C. Francioli, Grace Tiao, et al. 2020. “The Mutational Constraint Spectrum Quantified from Variation in 141,456 Humans.” Nature 581 (7809): 434–43. 10.1038/s41586-020-2308-7.

Katoh, Kazutaka, Kazuharu Misawa, Kei-ichi Kuma, and Takashi Miyata. 2002. “MAFFT: A Novel Method for Rapid Multiple Sequence Alignment Based on Fast Fourier Transform.” Nucleic Acids Research 30 (14): 3059–66. 10.1093/nar/gkf436.

Kępka, Alina, Agnieszka Ochocińska, Sylwia Chojnowska, et al. 2021. “Potential Role of L-Carnitine in Autism Spectrum Disorder.” Journal of Clinical Medicine 10 (6): 6. 10.3390/jcm10061202.

Kircher, Martin, Daniela M. Witten, Preti Jain, Brian J. O’Roak, Gregory M. Cooper, and Jay Shendure. 2014. “A General Framework for Estimating the Relative Pathogenicity of Human Genetic Variants.” Nature Genetics 46 (3): 310–15. 10.1038/ng.2892.

Kleinert, Philip, and Martin Kircher. 2022. “A Framework to Score the Effects of Structural Variants in Health and Disease.” Genome Research 32 (4): 766–77. 10.1101/gr.275995.121.

Koren, Sergey, Arang Rhie, Brian P. Walenz, et al. 2018. “De Novo Assembly of Haplotype-Resolved Genomes with Trio Binning.” Nature Biotechnology 36 (12): 1174–82. 10.1038/nbt.4277.

Kronenberg, Zev N., Edward J. Osborne, Kelsey R. Cone, et al. 2015. “Wham: Identifying Structural Variants of Biological Consequence.” PLOS Computational Biology 11 (12): e1004572. 10.1371/journal.pcbi.1004572.

Layer, Ryan M., Colby Chiang, Aaron R. Quinlan, and Ira M. Hall. 2014. “LUMPY: A Probabilistic Framework for Structural Variant Discovery.” Genome Biology 15 (6): R84. 10.1186/gb-2014-15-6-r84.

Leblond, Claire S., Freddy Cliquet, Coralie Carton, et al. 2019. “Both Rare and Common Genetic Variants Contribute to Autism in the Faroe Islands.” Npj Genomic Medicine 4 (1): 1. 10.1038/s41525-018-0075-2.

Li, Heng. 2021. “New Strategies to Improve Minimap2 Alignment Accuracy.” Bioinformatics 37 (23): 4572–74. 10.1093/bioinformatics/btab705.

Liao, Wen-Wei, Mobin Asri, Jana Ebler, et al. 2023. “A Draft Human Pangenome Reference.” Nature 617 (7960): 312–24. 10.1038/s41586-023-05896-x.

Lin, Jyun-Hong, Liang-Chi Chen, Shu-Chi Yu, and Yao-Ting Huang. 2022. “LongPhase: An Ultra-Fast Chromosome-Scale Phasing Algorithm for Small and Large Variants.” Bioinformatics 38 (7): 1816–22. 10.1093/bioinformatics/btac058.

Liu, Xiaoming, Chang Li, Chengcheng Mou, Yibo Dong, and Yicheng Tu. 2020. “dbNSFP v4: A Comprehensive Database of Transcript-Specific Functional Predictions and Annotations for Human Nonsynonymous and Splice-Site SNVs.” Genome Medicine 12 (1): 103. 10.1186/s13073-020-00803-9.

Logsdon, Glennis A., Peter Ebert, Peter A. Audano, et al. 2024. “Complex Genetic Variation in Nearly Complete Human Genomes.” Preprint, bioRxiv, September 25. 10.1101/2024.09.24.614721.

Logsdon, Glennis A., Allison N. Rozanski, Fedor Ryabov, et al. 2024. “The Variation and Evolution of Complete Human Centromeres.” Nature 629 (8010): 136–45. 10.1038/s41586-024-07278-3.

Logsdon, Glennis A., Mitchell R. Vollger, and Evan E. Eichler. 2020. “Long-Read Human Genome Sequencing and Its Applications.” Nature Reviews Genetics 21 (10): 597–614. 10.1038/s41576-020-0236-x.

Manolio, Teri A., Francis S. Collins, Nancy J. Cox, et al. 2009. “Finding the Missing Heritability of Complex Diseases.” Nature 461 (7265): 747–53. 10.1038/nature08494.

Mastrorosa, Francesco Kumara, Danny E. Miller, and Evan E. Eichler. 2023. “Applications of Long-Read Sequencing to Mendelian Genetics.” Genome Medicine 15 (1): 42. 10.1186/s13073-023-01194-3.

McLaren, William, Laurent Gil, Sarah E. Hunt, et al. 2016. “The Ensembl Variant Effect Predictor.” Genome Biology 17 (1): 122. 10.1186/s13059-016-0974-4.

Miller, Danny E., Arvis Sulovari, Tianyun Wang, et al. 2021. “Targeted Long-Read Sequencing Identifies Missing Disease-Causing Variation.” The American Journal of Human Genetics 108 (8): 1436–49. 10.1016/j.ajhg.2021.06.006.

Noyes, Michelle D., William T. Harvey, David Porubsky, et al. 2022. “Familial Long-Read Sequencing Increases Yield of *de Novo* Mutations.” The American Journal of Human Genetics 109 (4): 631–46. 10.1016/j.ajhg.2022.02.014.

Noyes, Michelle D., Yang Sui, Youngjun Kwon, et al. 2025. “Long-Read Sequencing of Trios Reveals Increased Germline and Postzygotic Mutation Rates in Repetitive DNA.” Preprint, bioRxiv, July 19. 10.1101/2025.07.18.665621.

Olivucci, Giulia, Emanuela Iovino, Giovanni Innella, Daniela Turchetti, Tommaso Pippucci, and Pamela Magini. 2024. “Long Read Sequencing on Its Way to the Routine Diagnostics of Genetic Diseases.” Frontiers in Genetics 15 (March). 10.3389/fgene.2024.1374860.

Paschal, Cate R., Miranda P. G. Zalusky, Anita E. Beck, et al. 2025. “Concordance of Whole-Genome Long-Read Sequencing with Standard Clinical Testing for Prader-Willi and Angelman Syndromes.” The Journal of Molecular Diagnostics 27 (3): 166–76. 10.1016/j.jmoldx.2024.12.003.

Pauper, Marc, Erdi Kucuk, Aaron M. Wenger, et al. 2021. “Long-Read Trio Sequencing of Individuals with Unsolved Intellectual Disability.” European Journal of Human Genetics 29 (4): 637–48. 10.1038/s41431-020-00770-0.

Pedersen, Brent S., Preetida J. Bhetariya, Joe Brown, et al. 2020. “Somalier: Rapid Relatedness Estimation for Cancer and Germline Studies Using Efficient Genome Sketches.” Genome Medicine 12 (1): 62. 10.1186/s13073-020-00761-2.

Perez, Gerardo, Galt P Barber, Anna Benet-Pages, et al. 2025. “The UCSC Genome Browser Database: 2025 Update.” Nucleic Acids Research 53 (D1): D1243–49. 10.1093/nar/gkae974.

Poplin, Ryan, Pi-Chuan Chang, David Alexander, et al. 2018. “A Universal SNP and Small-Indel Variant Caller Using Deep Neural Networks.” Nature Biotechnology 36 (10): 983–87. 10.1038/nbt.4235.

Poplin, Ryan, Valentin Ruano-Rubio, Mark A. DePristo, et al. 2018. “Scaling Accurate Genetic Variant Discovery to Tens of Thousands of Samples.” Preprint, bioRxiv, July 24. 10.1101/201178.

Porubsky, David, Harriet Dashnow, Thomas A. Sasani, et al. 2025. “Human de Novo Mutation Rates from a Four-Generation Pedigree Reference.” Nature, April 23, 1–10. 10.1038/s41586-025-08922-2.

Porubsky, David, Xavi Guitart, DongAhn Yoo, Philip C Dishuck, William T Harvey, and Evan E Eichler. 2025. “SVbyEye: A Visual Tool to Characterize Structural Variation among Whole-Genome Assemblies.” Bioinformatics 41 (6): btaf332. 10.1093/bioinformatics/btaf332.

Rausch, Tobias, Thomas Zichner, Andreas Schlattl, Adrian M. Stütz, Vladimir Benes, and Jan O. Korbel. 2012. “DELLY: Structural Variant Discovery by Integrated Paired-End and Split-Read Analysis.” Bioinformatics 28 (18): i333–39. 10.1093/bioinformatics/bts378.

Reid, Kyle M., and Guy C. Brown. 2023. “LRPAP1 Is Released from Activated Microglia and Inhibits Microglial Phagocytosis and Amyloid Beta Aggregation.” Frontiers in Immunology 14 (November). 10.3389/fimmu.2023.1286474.

Rhie, Arang, Sergey Nurk, Monika Cechova, et al. 2023. “The Complete Sequence of a Human Y Chromosome.” Nature 621 (7978): 344–54. 10.1038/s41586-023-06457-y.

Rhie, Arang, Brian P. Walenz, Sergey Koren, and Adam M. Phillippy. 2020. “Merqury: Reference-Free Quality, Completeness, and Phasing Assessment for Genome Assemblies.” Genome Biology 21 (1): 245. 10.1186/s13059-020-02134-9.

Rimmer, Andy, Hang Phan, Iain Mathieson, et al. 2014. “Integrating Mapping-, Assembly- and Haplotype-Based Approaches for Calling Variants in Clinical Sequencing Applications.” Nature Genetics 46 (8): 912–18. 10.1038/ng.3036.

Robinson, James T., Helga Thorvaldsdóttir, Aaron M. Wenger, Ahmet Zehir, and Jill P. Mesirov. 2017. “Variant Review with the Integrative Genomics Viewer.” Cancer Research 77 (21): e31–34. 10.1158/0008-5472.CAN-17-0337.

Roller, Eric, Sergii Ivakhno, Steve Lee, Thomas Royce, and Stephen Tanner. 2016. “Canvas: Versatile and Scalable Detection of Copy Number Variants.” Bioinformatics 32 (15): 2375–77. 10.1093/bioinformatics/btw163.

Rosa e Silva, Ivan, Juliana Helena Costa Smetana, and Juliana Ferreira de Oliveira. 2024. “A Comprehensive Review on DDX3X Liquid Phase Condensation in Health and Neurodevelopmental Disorders.” International Journal of Biological Macromolecules 259 (February): 129330. 10.1016/j.ijbiomac.2024.129330.

Sanchis-Juan, Alba, Karyn Megy, Jonathan Stephens, et al. 2023. “Genome Sequencing and Comprehensive Rare-Variant Analysis of 465 Families with Neurodevelopmental Disorders.” The American Journal of Human Genetics 110 (8): 1343–55. 10.1016/j.ajhg.2023.07.007.

Sanders, Stephan J., A. Gulhan Ercan-Sencicek, Vanessa Hus, et al. 2011. “Multiple Recurrent De Novo CNVs, Including Duplications of the 7q11.23 Williams Syndrome Region, Are Strongly Associated with Autism.” Neuron 70 (5): 863–85. 10.1016/j.neuron.2011.05.002.

Satterstrom, F. Kyle, Jack A. Kosmicki, Jiebiao Wang, et al. 2020. “Large-Scale Exome Sequencing Study Implicates Both Developmental and Functional Changes in the Neurobiology of Autism.” Cell 180 (3): 568–584.e23. 10.1016/j.cell.2019.12.036.

Scott, Alexandra J., Colby Chiang, and Ira M. Hall. 2021. “Structural Variants Are a Major Source of Gene Expression Differences in Humans and Often Affect Multiple Nearby Genes.” Genome Research 31 (12): 2249–57. 10.1101/gr.275488.121.

Sebat, Jonathan, B. Lakshmi, Dheeraj Malhotra, et al. 2007. “Strong Association of De Novo Copy Number Mutations with Autism.” Science 316 (5823): 445–49. 10.1126/science.1138659.

Sierra, Adriana Y., Esther Gratacós, Patricia Carrasco, et al. 2008. “CPT1c Is Localized in Endoplasmic Reticulum of Neurons and Has Carnitine Palmitoyltransferase Activity*.” Journal of Biological Chemistry 283 (11): 6878–85. 10.1074/jbc.M707965200.

Smolka, Moritz, Luis F. Paulin, Christopher M. Grochowski, et al. 2024. “Detection of Mosaic and Population-Level Structural Variants with Sniffles2.” Nature Biotechnology 42 (10): 1571–80. 10.1038/s41587-023-02024-y.

Stessman, Holly A. F., Marjolein H. Willemsen, Michaela Fenckova, et al. 2016. “Disruption of POGZ Is Associated with Intellectual Disability and Autism Spectrum Disorders.” The American Journal of Human Genetics 98 (3): 541–52. 10.1016/j.ajhg.2016.02.004.

Sudmant, Peter H., Swapan Mallick, Bradley J. Nelson, et al. 2015. “Global Diversity, Population Stratification, and Selection of Human Copy-Number Variation.” Science 349 (6253): aab3761. 10.1126/science.aab3761.

Sudmant, Peter H., Tobias Rausch, Eugene J. Gardner, et al. 2015. “An Integrated Map of Structural Variation in 2,504 Human Genomes.” Nature 526 (7571): 75–81. 10.1038/nature15394.

Tillotson, Rebekah, and Adrian Bird. 2020. “The Molecular Basis of MeCP2 Function in the Brain.” *Journal of Molecular Biology*, Reading DNA Modifications, vol. 432 (6): 1602–23. 10.1016/j.jmb.2019.10.004.

Trost, Brett, Worrawat Engchuan, Charlotte M. Nguyen, et al. 2020. “Genome-Wide Detection of Tandem DNA Repeats That Are Expanded in Autism.” Nature 586 (7827): 80–86. 10.1038/s41586-020-2579-z.

Turner, Tychele N., Bradley P. Coe, Diane E. Dickel, et al. 2017. “Genomic Patterns of De Novo Mutation in Simplex Autism.” Cell 171 (3): 710–722.e12. 10.1016/j.cell.2017.08.047.

Wang, Tianyun, Chang N. Kim, Trygve E. Bakken, et al. 2022. “Integrated Gene Analyses of de Novo Variants from 46,612 Trios with Autism and Developmental Disorders.” Proceedings of the National Academy of Sciences 119 (46): e2203491119. 10.1073/pnas.2203491119.

Waterhouse, Andrew M., James B. Procter, David M. A. Martin, Michèle Clamp, and Geoffrey J. Barton. 2009. “Jalview Version 2—a Multiple Sequence Alignment Editor and Analysis Workbench.” Bioinformatics 25 (9): 1189–91. 10.1093/bioinformatics/btp033.

Wilfert, Amy B., Tychele N. Turner, Shwetha C. Murali, et al. 2021. “Recent Ultra-Rare Inherited Variants Implicate New Autism Candidate Risk Genes.” Nature Genetics 53 (8): 1125–34. 10.1038/s41588-021-00899-8.

Wojcik, Monica H., Chloe M. Reuter, Shruti Marwaha, et al. 2023. “Beyond the Exome: What’s next in Diagnostic Testing for Mendelian Conditions.” The American Journal of Human Genetics 110 (8): 1229–48. 10.1016/j.ajhg.2023.06.009.

Wood, Derrick E., Jennifer Lu, and Ben Langmead. 2019. “Improved Metagenomic Analysis with Kraken 2.” Genome Biology 20 (1): 257. 10.1186/s13059-019-1891-0.

Yoon, Seungtai, Adriana Munoz, Boris Yamrom, et al. 2021. “Rates of Contributory de Novo Mutation in High and Low-Risk Autism Families.” Communications Biology 4 (1): 1026. 10.1038/s42003-021-02533-z.

Zaghlula, Manar, Daniel G. Glaze, Gregory M. Enns, Lorraine Potocki, Aloysia L. Schwabe, and Bernhard Suter. 2018. “Current Clinical Evidence Does Not Support a Link between TBL1XR1 and Rett Syndrome: Description of One Patient with Rett Features and a Novel Mutation in TBL1XR1, and a Review of TBL1XR1 Phenotypes.” American Journal of Medical Genetics Part A 176 (7): 1683–87. 10.1002/ajmg.a.38689.

Zeidan, Jinan, Eric Fombonne, Julie Scorah, et al. 2022. “Global Prevalence of Autism: A Systematic Review Update.” Autism Research 15 (5): 778–90. 10.1002/aur.2696.

Zhang, Fan, Matthew Flickinger, Sarah A. Gagliano Taliun, et al. 2020. “Ancestry-Agnostic Estimation of DNA Sample Contamination from Sequence Reads.” Genome Research 30 (2): 185–94. 10.1101/gr.246934.118.

Zhao, Xuefang, Ryan L. Collins, Wan-Ping Lee, et al. 2021. “Expectations and Blind Spots for Structural Variation Detection from Long-Read Assemblies and Short-Read Genome Sequencing Technologies.” The American Journal of Human Genetics 108 (5): 919–28. 10.1016/j.ajhg.2021.03.014.

Zheng, Zhenxian, Shumin Li, Junhao Su, Amy Wing-Sze Leung, Tak-Wah Lam, and Ruibang Luo. 2022. “Symphonizing Pileup and Full-Alignment for Deep Learning-Based Long-Read Variant Calling.” Nature Computational Science 2 (12): 797–803. 10.1038/s43588-022-00387-x.

Zhou, Xueya, Pamela Feliciano, Chang Shu, et al. 2022. “Integrating de Novo and Inherited Variants in 42,607 Autism Cases Identifies Mutations in New Moderate-Risk Genes.” Nature Genetics 54 (9): 1305–19. 10.1038/s41588-022-01148-2.

