## Supplemental Note and Figures for "Pangenome discovery of missing autism variants"

### Table of Contents

Supplemental Note
Supplemental Figures S1-S10
Supplemental Tables S1-S3

### Supplemental Note

#### Human Pangenome Reference Consortium Release 2 Authors

Ahmad Abou Tayoun[1,2], Derek Albracht[3], Jamie Allen[4], Alawi A. Alsheikh-Ali[5], Casey Andrews[6], Dmitry Antipov[7], Lucinda Antonacci-Fulton[3], Mobin Asri[8], Marcelo Ayllon[9], Jennifer R. Balacco[10], Edward A Belter Jr[3], Halle D. Bender[8], Andrew P. Blair[8], Silvia Buonaiuto[11], Davide Bolognini[12], Katherine E. Bonini[13], Christina Boucher[14], Guillaume Bourque[15,16,17], Shuo Cao[11], Andrew Carroll[18], Ann M. Mc Cartney[8], Monika Cechova[8], Pi-Chuan Chang[18], Xian Chang[8], Jitender Cheema[4], Haoyu Cheng[19], Claudio Ciofi[20], Sarah Cody[3], Vincenza Colonna[11], Holland C. Conwell[21], Robert Cook-Deegan[22], Mark Diekhans[8], Maria Angela Diroma[20], Daniel Doerr[23,24,25], Zheng Dong[6], Richard Durbin[26,27], Jana Ebler[23,28], Evan E Eichler[9,29], Jordan M. Eizenga[8], Parsa Eskandar[8], Eddie Ferro[14], Anna-Sophie Fiston-Lavier[30,31], Sarah M. Ford[21], Willard W. Ford[32], Giulio Formenti[10], Adam Frankish[4], Mallory A. Freeberg[4], Qichen Fu[6], Stephanie M. Fullerton[33], Robert S. Fulton[3], Yan Gao[34], Gage H. Garcia[9], Obed A. Garcia[35], Joshua M.V. Gardner[8], Shilpa Garg[36], Erik Garrison[11], Nanibaa' A. Garrison[37,38,39], John Garza[3], Margarita Geleta[8], Mohammadmersad Ghorbani[40], Sky Gomez[8], Tina Graves-Lindsay[3], Richard E. Green[21], Cristian Groza[41], Andrea Guarracino[11], Melissa Gymrek[32], Leanne Haggerty[4], Ira M Hall[42,43], Nancy F. Hansen[7], Mohammad Amiruddin Hashmi[5], Maximilian Haeussler[8], David Haussler[8], Prajna Hebbar[8], Peter Heringer[23,24,25], Glenn Hickey[8], Todd L. Hillaker[8], S. Nakib Hossain[4], Neng Huang[34,44], Sarah E Hunt[4], Toby Hunt[4], Alexander G. Ioannidis[8], Nafiseh Jafarzadeh[8], Nivesh Jain[10], Erich D. Jarvis[10,29], Juan Jiang[6], Jonathan LoTempio Jr[45], Eimear E. Kenny[13], Juhyun Kim[7], Bonhwang Koo[10], Sergey Koren[7], Milinn Kremitzki[3,6], Ben Langmead[46], Xiaoyu Zhuo[6], Heather A. Lawson[6], Daofeng Li[6], Heng Li[34,44], Wen-Wei Liao[42,43], Jiadong Lin[9], Tianjie Liu[6], Glennis A. Logsdon[45], Ryan Lorig-Roach[8], Hailey Loucks[8], Jane E Loveland[4], Jianguo Lu[47], Shuangjia Lu[42,43], Julian K. Lucas[8], Juan F. Macias-Velasco[3,6,48], Maximillian G. Marin[34], Franco L. Marsico[11], Kateryna D. Makova[49], Christopher Markovic[6], Tobias Marschall[23,28], Fergal J Martin[4], Mira Mastoras[8], Capucine Mayoud[30], Brandy McNulty[8], Jack A. Medico[10], Julian M. Menendez[8], Karen H. Miga[8], Anna Minkina[50], Matthew W. Mitchell[51], Saswat K. Mohanty[52], Younes Mokrab[40,53,54], Jean Monlong[55], Shabir Moosa[40], Avelina Moreno-Ochando[56,57], Shinichi Morishita[58], Jonathan M. Mudge[4], Katherine M. Munson[9], Njagi Mwaniki[59], Nasna Nassir[5], Chiara Natali[20], Shloka Negi[8], Lingbin Ni[9], Adam M. Novak[8], Chie Owa[58], Sadye Paez[10], Benedict Paten[8], Hiram Clawson[8], Clelia Peano[12,60], Adam M. Phillippy[7], Brandon D. Pickett[7], Laura Pignata[11], Nadia Pisanti[59], David Porubsky[9], Pjotr Prins[11], Anandi Radhakrishnan[8], Brian J. Raney[8], Mikko Rautiainen[61],

Alessandro Raveane[12], Luyao Ren[9,29], Arang Rhie[7], Farnaz Salehi[11], Samuel Sacco[21], Michael C. Schatz[46,62], Laura B. Scheinfeldt[51], Aarushi Sehgal[32], William E. Seligmann[21], Mahsa Shabani[63], Kishwar Shafin[18], Shadi Shahatit[30], Ruhollah Shemirani[13], Vikram S. Shivakumar[46], Swati Sinha[4], Jouni Sirén[8], Linnéa Smeds[52], Steven J. Solar[7], Marco Sollitto[10,20], Nicole Soranzo[12,26,27], Andrew B Stergachis[9,50], Marie-Marthe Suner[4], Yoshihiko Suzuki[58], Arda Söylev[23,28], Jack AS Tierney[4], Chad Tomlinson[3], Francesca Floriana Tricomi[4], Mohammed Uddin[5,64], Matteo Tommaso Ungaro[21,65], Rahul Varki[14], Flavia Villani[11], Mitchell R. Vollger[50], Brian P. Walenz[7], Charles Wang[66], Lisa E. Wang[13], Ting Wang[3,6,48], Aaron M. Wenger[67], Conor V. Whelan[10], Zilan Xin[6], Zheng Xu[6], Kai Ye[68], DongAhn Yoo[9], Wenjin Zhang[6], Ying Zhou[34], Ivo Violich[8], Giulia Zunino[12]

### Affiliations

1. Center for Genomic Discovery, Mohammed Bin Rashid University, Dubai Health, UAE
2. Dubai Health Genomic Medicine Center, Dubai Health, UAE
3. McDonnell Genome Institute, Washington University School of Medicine, St. Louis, MO 63108, USA
4. European Molecular Biology Laboratory, European Bioinformatics Institute (EMBL-EBI), Wellcome Genome Campus, Hinxton, Cambridge CB10 1SD, UK
5. Center for Applied and Translational Genomics (CATG), Mohammed Bin Rashid University of Medicine and Health Sciences, Dubai, United Arab Emirates
6. Department of Genetics, Washington University School of Medicine, St. Louis, MO 63110, USA
7. Genome Informatics Section, Center for Genomics and Data Science Research, National Human Genome Research Institute, National Institutes of Health, Bethesda, MD 20894, USA
8. UC Santa Cruz Genomics Institute, University of California, Santa Cruz, 2300 Delaware Avenue, Santa Cruz, CA 95060, USA
9. Department of Genome Sciences, University of Washington School of Medicine, Seattle, WA 98195, USA
10. The Vertebrate Genome Laboratory, The Rockefeller University, 1230 York Ave, NY 10065, USA
11. Department of Genetics, Genomics and Informatics, University of Tennessee Health Science Center, 71 S Manassas St, Memphis, TN 38163, USA
12. Human Technopole, Milan, Italy
13. Institute for Genomic Health, Icahn School of Medicine at Mount Sinai, New York, NY 10029, USA
14. Department of Computer and Information Science and Engineering, University of Florida, Gainesville, FL 32611, USA
15. Canadian Center for Computational Genomics, McGill University, Montréal, QC, Canada
16. Department of Human Genetics, McGill University, Montréal, QC, Canada
17. Victor Phillip Dahdaleh Institute of Genomic Medicine, Montréal, QC, Canada
18. Google LLC, 1600 Amphitheatre Pkwy, Mountain View, CA 94043, USA
19. Department of Biomedical Informatics and Data Science, Yale School of Medicine, New Haven, CT 06510, USA
20. University of Florence, Department of Biology, Via Madonna del Piano, 6, Sesto

Fiorentino (FI) 50019, Italy
21. Department of Ecology and Evolutionary Biology, University of California, Santa Cruz, Santa Cruz, CA 95064, USA
22. Arizona State University, Consortium for Science, Policy & Outcomes, 1800 I St NW, Washington, DC 20006, USA
23. Center for Digital Medicine, Heinrich Heine University Düsseldorf, Germany
24. Department for Endocrinology and Diabetology, Medical Faculty and University Hospital Düsseldorf, Heinrich Heine University Düsseldorf, Germany
25. German Diabetes Center (DDZ), Leibniz Institute for Diabetes Research, Düsseldorf, Germany
26. University of Cambridge, Cambridge, UK
27. Wellcome Sanger Institute, Genome Campus, Hinxton, CB10 1HH, UK
28. Institute for Medical Biometry and Bioinformatics, Medical Faculty and University Hospital Düsseldorf, Heinrich Heine University, Düsseldorf, Germany
29. Howard Hughes Medical Institute, Chevy Chase, MD 20815, USA
30. ISEM, Univ Montpellier, CNRS, IRD, Montpellier, France
31. Institut Universitaire de France, Paris, France
32. Department of Computer Science and Engineering, University of California San Diego, La Jolla, CA 92093, USA
33. Department of Bioethics & Humanities, University of Washington School of Medicine, Seattle, WA 98195, USA
34. Department of Data Science, Dana-Farber Cancer Institute, MA 02215, USA
35. Department of Anthropology, University of Kansas, KS 66044 USA
36. University of Manchester, Manchester M13 9PL, UK
37. Traditional, ancestral and unceded territory of the Gabrielino/Tongva peoples, Institute for Society & Genetics, University of California, Los Angeles, Los Angeles, CA 90095, USA
38. Traditional, ancestral and unceded territory of the Gabrielino/Tongva peoples, Institute for Precision Health, David Geffen School of Medicine, University of California, Los Angeles, Los Angeles, CA 90095, USA
39. Traditional, ancestral and unceded territory of the Gabrielino/Tongva peoples, Division of General Internal Medicine & Health Services Research, David Geffen School of Medicine, University of California, Los Angeles, Los Angeles, CA 90095, USA
40. Medical and Population Genomics Lab, Sidra Medicine, Doha, Qatar
41. Montreal Heart Institute, Montreal, Quebec, Canada
42. Center for Genomic Health, Yale University School of Medicine, New Haven, CT 06510, USA
43. Department of Genetics, Yale University School of Medicine, New Haven, CT 06510, USA
44. Department of Biomedical Informatics, Harvard Medical School, MA 02115, USA
45. Department of Genetics, Epigenetics Institute, Perelman School of Medicine, University of Pennsylvania, Philadelphia, PA 19104, USA
46. Department of Computer Science, Johns Hopkins University, Baltimore, MD 21218, USA
47. Sun Yat-sen University, No. 135, Xingang Xi Road, Guangzhou, China
48. Edison Family Center for Genome Sciences & Systems Biology, Washington University

School of Medicine, St. Louis, MO 63110, USA
49. Center for Medical Genomics, Penn State University, University Park, PA 16802, USA
50. Division of Medical Genetics, Department of Medicine, University of Washington School of Medicine, Seattle, WA 98195, USA
51. Coriell Institute for Medical Research, Camden, NJ 08103, USA
52. Department of Biology, Penn State University, University Park, PA 16802, USA
53. Department of Biomedical Science, College of Health Sciences, Qatar University, Doha, Qatar
54. Department of Genetic Medicine, Weill Cornell Medicine-Qatar, Doha, Qatar
55. IRSD - Digestive Health Research Institute, University of Toulouse, INSERM, INRAE, ENVT, UPS, Toulouse, France
56. MATCH biosystems, S.L., Spain
57. Universidad Miguel Hernández de Elche, Spain
58. Department of Computational Biology and Medical Sciences, The University of Tokyo, Kashiwa, Chiba 277-8561, Japan
59. University of Pisa, Pisa, Italy
60. Institute of Genetics and Biomedical Research, UoS of Milan, National Research Council, Milan, Italy
61. Institute for Molecular Medicine Finland, Helsinki Institute of Life Science, University
62. Department of Biology, Johns Hopkins University, Baltimore, MD 21218, USA
63. University of Amsterdam, Amsterdam, Netherlands
64. GenomeArc Inc, Mississauga, ON, Canada
65. Department of Biology and Biotechnologies "Charles Darwin", University of Rome "La Sapienza", 00185 Rome, Italy
66. Center for Genomics, Loma Linda University School of Medicine, Loma Linda, CA 92350, USA
67. PacBio, 1305 O'Brien Drive, Menlo Park, CA 94025, USA
68. The first affiliated hospital of Xi'an Jiaotong University, Xi'an Jiaotong University, Xi'an, Shaanxi, 710049, China

### Funding Statement

We would like to acknowledge the National Genome Research Institute (NHGRI) for funding the following grants supporting the creation of the human pangenome reference: U41HG010972, U01HG010971, U01HG013760, U01HG013755, U01HG013748, U01HG013744, R01HG011274, and the Human Pangenome Reference Consortium (BioProject ID: PRJNA730823).

### Supplemental Figures

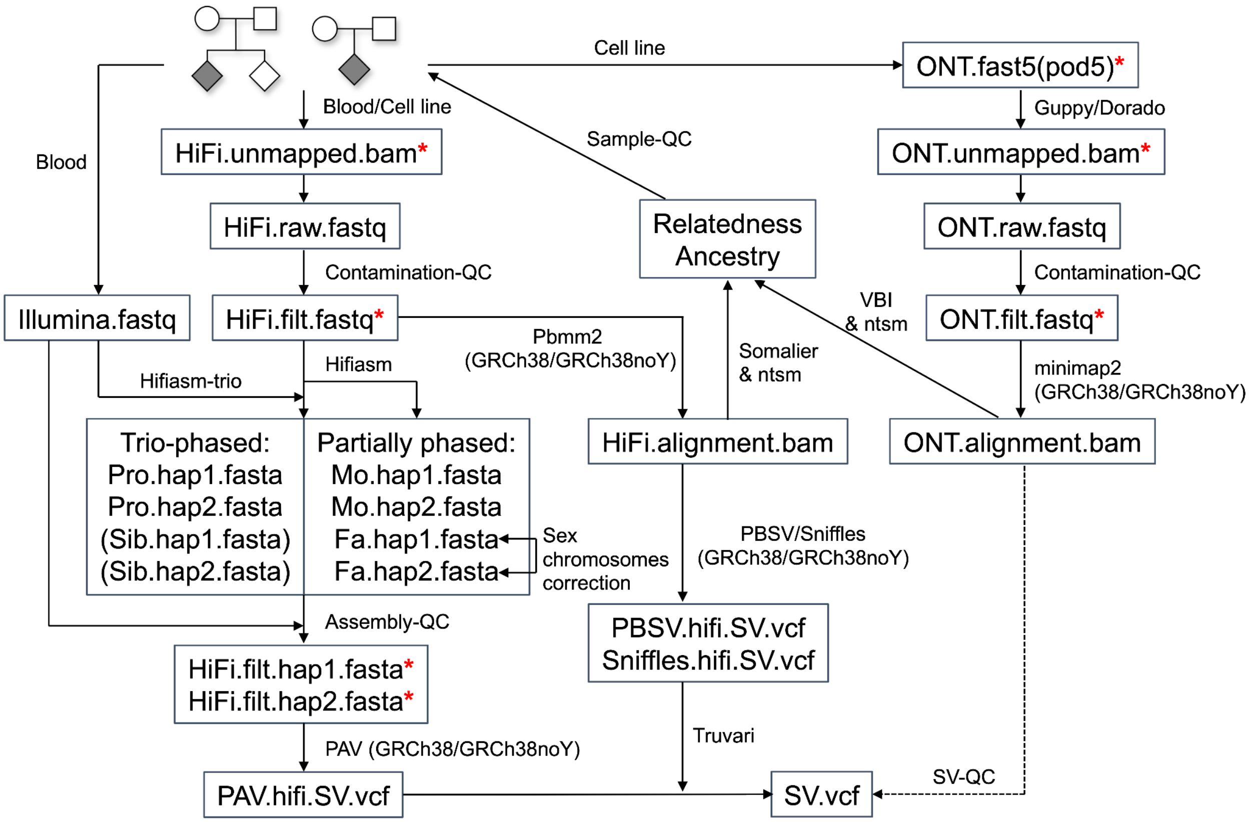


**Figure S1. LRS data QC and processing.**

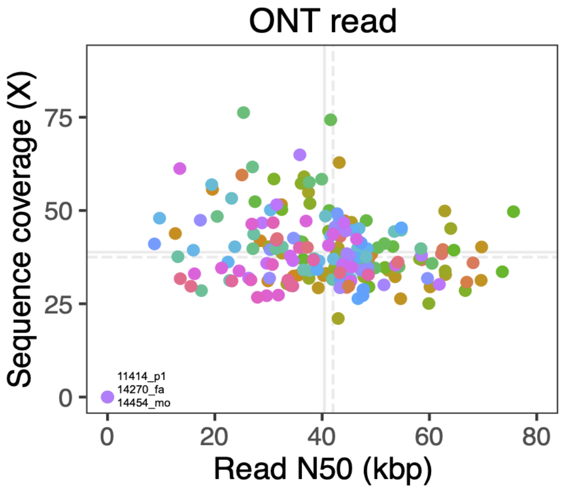


**Figure S2. ONT sequencing coverage and reads N50 in 51 unsolved families.** 186 of 189 individuals have ONT data available (members of the same family are color coded). Solid lines represent mean values, while dashed lines indicate median values.

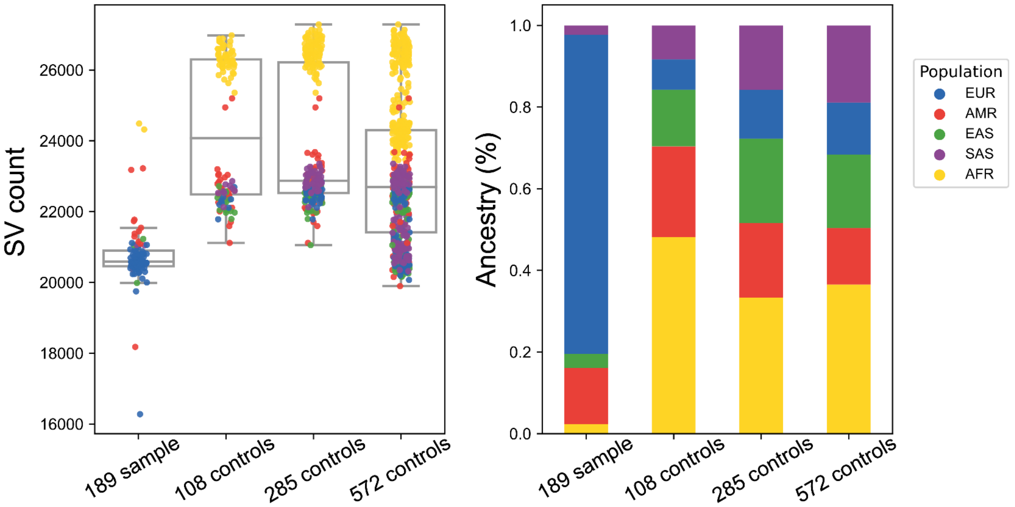


**Figure S3. Validated SV count in long-read pangenome controls and 51 ASD families.** The 108 control sets were from HPRC and HGSVC. The 285 control sets contain an additional 177 HPRC individuals. The 572 control sets contain additional 287 1KG individuals. The ancestry of 189 individuals was predicted by Somalier (Pedersen et al. 2020) using HiFi alignments.

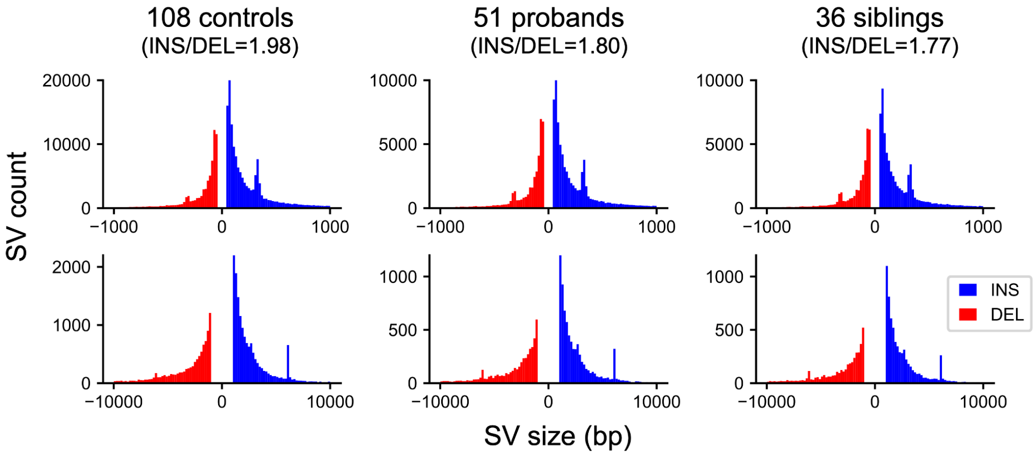


**Figure S4. Size distribution (bp) of nonredundant SVs in 108 controls, 51 probands and 36 siblings.** The ratio of INS to DEL is marked in the parentheses.

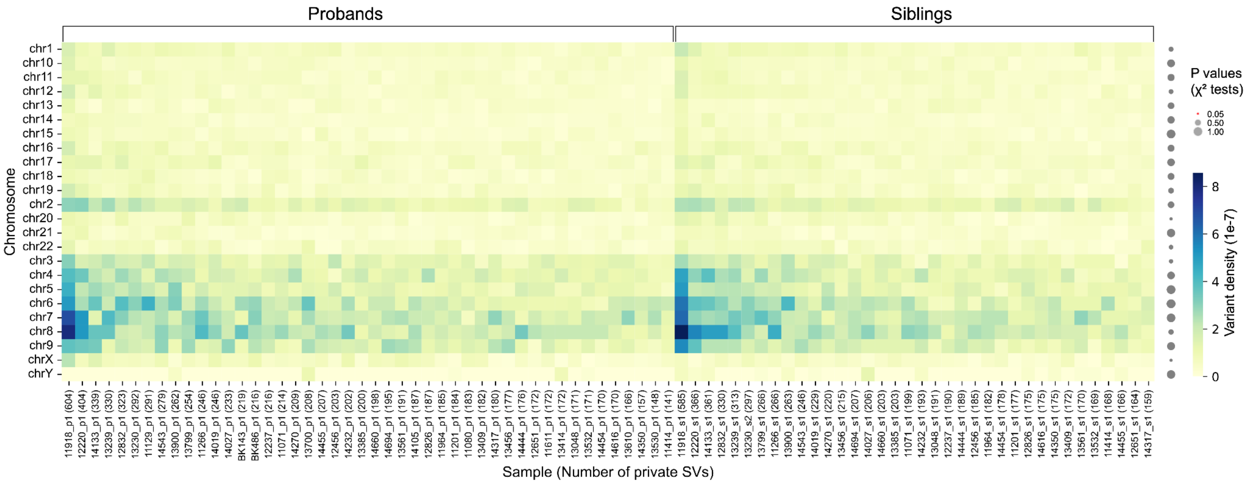


**Figure S5. Density heatmap (per chromosome) of private SVs.**

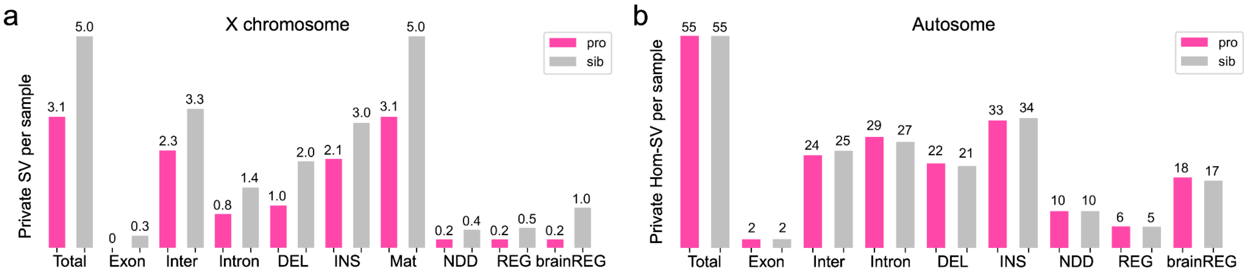


**Figure S6. Private SVs on X per male (a) and private homozygous SV on autosomes per child (b) as a function of their categories, genomic location, SV types.**

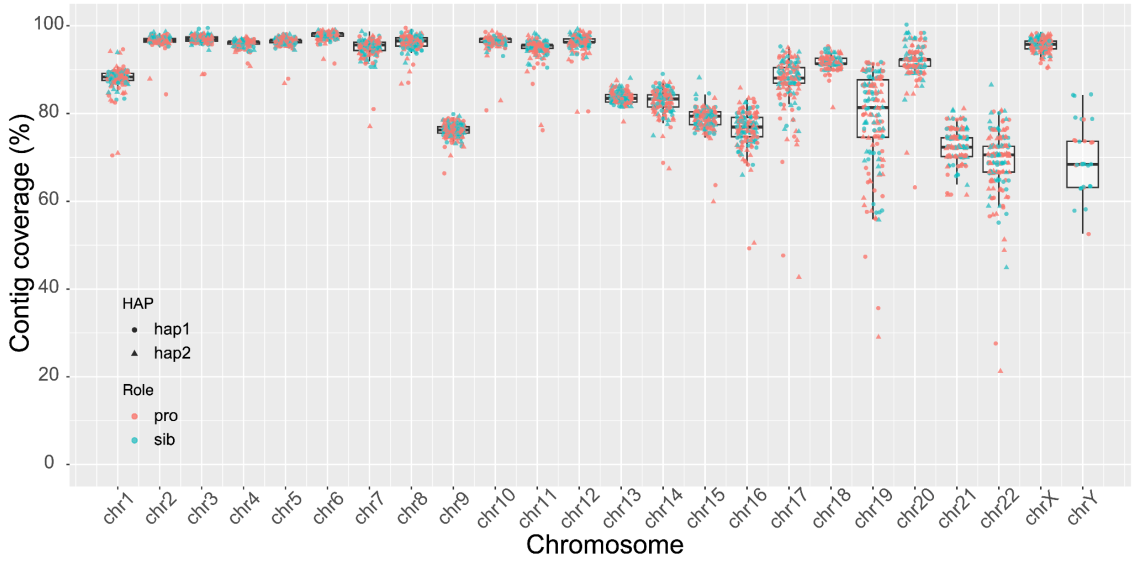


**Figure S7. Contig coverage relative to the T2T-CHM13v2.0 reference.** Each reference chromosome was divided into 1 Mbp windows and identified those covered by contigs that aligned to ≥95% of the window sequence with no more than three overlapping contigs. The coverage percentage was calculated as the number of qualified windows divided by the total number of windows, representing coverage relative to the reference chromosomes.

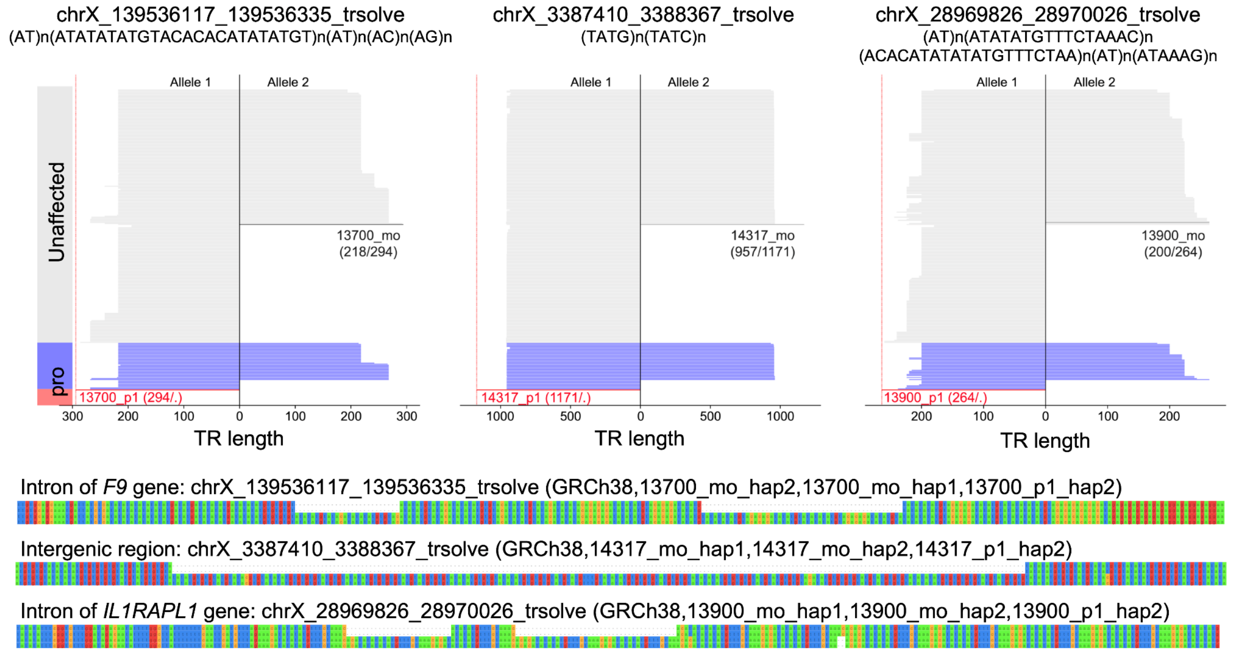


**Figure S8. Outlier TR expansions transmitted from mothers to male probands.** Allele lengths (generated by TRGT (Dolzhenko et al. 2024)) of three TR catalogs (Porubsky, Dashnow, et al. 2025) are shown in gray for unaffected individuals (108 controls and 128 unaffected samples) and in blue for affected probands. The three outlier expansions are highlighted in red and dark gray. Transmission patterns from maternal haplotypes to male probands are illustrated using multiple sequence alignments (MSAs), with comparisons to the GRCh38 reference. Note that females, with two X chromosomes, display bidirectional TR lengths (both alleles), whereas males, with only one X chromosome, show unidirectional TR length from a single allele.

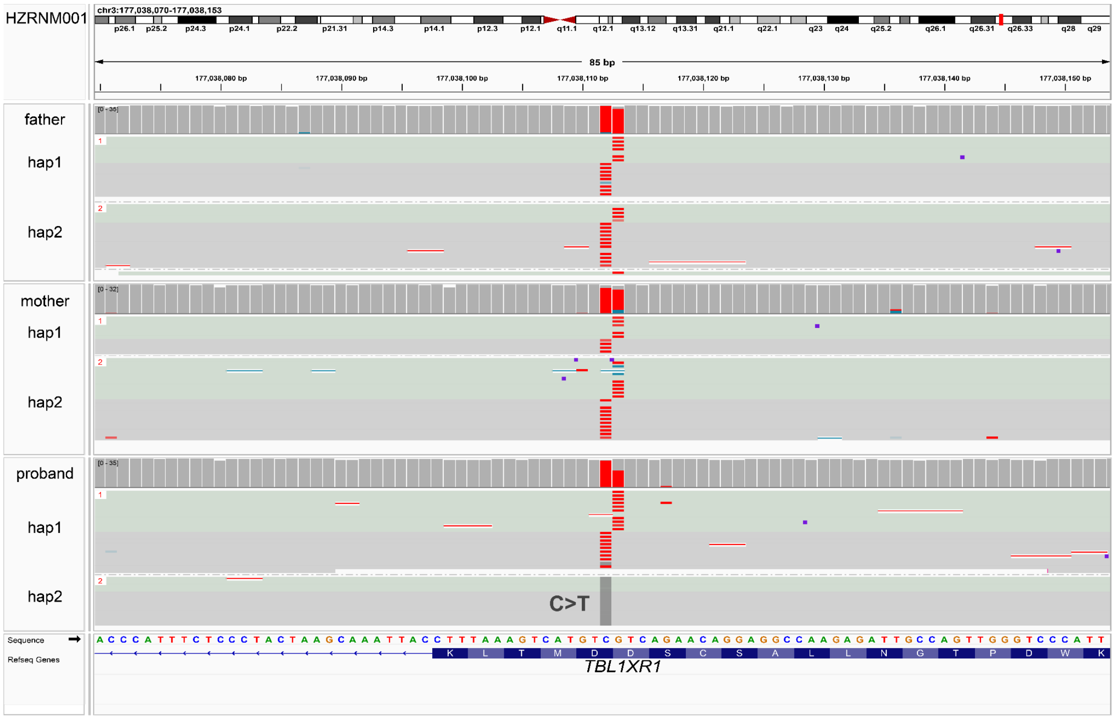


**Figure S9. Methylation changes at the CpG site induced by a DNM in *TBL1XR1*.**

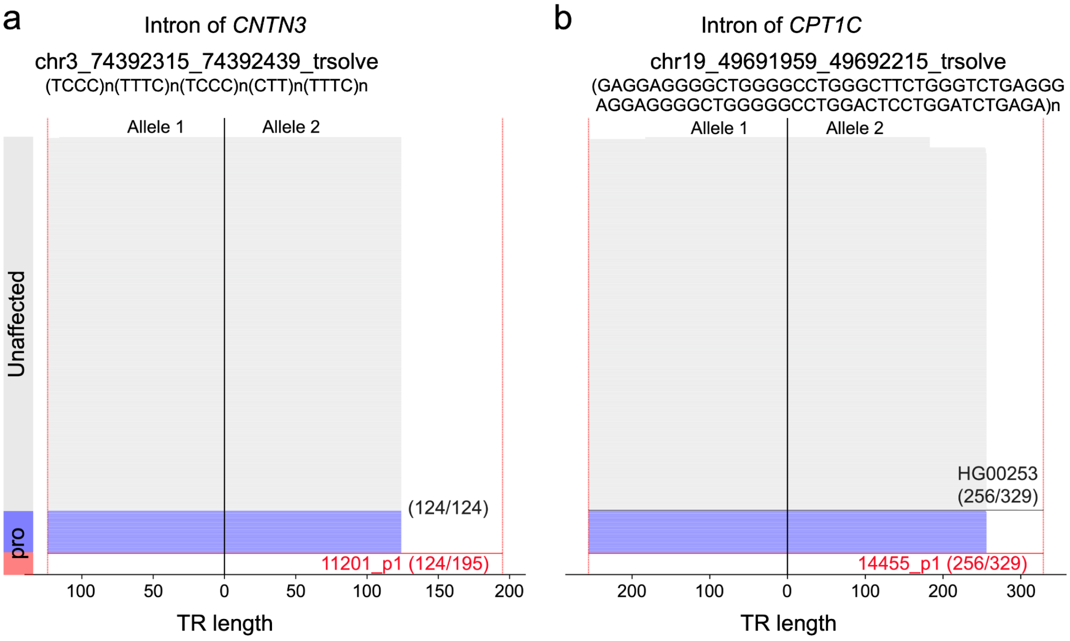


**Figure S10. *De novo* candidates validated in larger control sets.** The two *de novo* TR INS candidates were further examined in the 270 controls using TRGT with HiFi alignments. The 71 bp INS (**a**, red) in the TR remains a distinct outlier even when compared to an expanded control set. However, the 73 bp INS (**b**, red) were seen in one control. TR length of unaffected individuals and probands were shown in gray and blue, respectively.

### Supplemental Tables

**Dataset S1. LRS of unsolved autism families.**
**Dataset S2. Collapsed SVs.**
**Dataset S3. Table of pathogenic and candidate variants.**
